# The Orphanet Nomenclature and Classification of rare diseases: a standard terminology for improved patient recognition and data interoperability

**DOI:** 10.1101/2025.08.10.25333394

**Authors:** Caterina Lucano, David Lagorce, Annie Olry, Houda Ali, Valérie Lanneau, Mickael De Carvalho, Aysegul Dilsizoglu Senol, Marta Fructuoso, Emilie Gaillard, Marie-Cécile Gaillard, Seed Mihic, Mariana Tannoury, Florence Sauvage, Charlotte Rodwell, Sylvie Maiella, Marc Hanauer, Ana Rath

## Abstract

**Background:** Although individually uncommon, rare diseases (RD) collectively affect an estimated 329-624 million people worldwide There are over 6,500 known RD, 85% of which affect fewer than 1 person per million. As a result, the critical amount of data necessary to improve knowledge, care, and treatment can only be achieved through cumulative data collection across different countries in a standardized manner. However, RD are under-represented in medical terminologies and classification systems, hindering data sharing, interoperability, and public health monitoring.

**Objective:** The aim of this paper is to present the Orphanet Nomenclature and Classification of RD. We detail its content as well as its production and update methodology, and an overview of its mappings to other semantic resources. In addition, this work provides a clear and up-to-date count of RD based on the consensus operational definition of RD, and it details the distribution of RD by medical domain.

**Methods:** The Orphanet Nomenclature of RD is a multilingual standardized system composed of clinical entities, each defined by a unique and time-stable ORPHAcode, a preferred term, synonyms, a classification level, and a textual definition. This nomenclature is structured into three classification levels organized within a multi-hierarchical and multi-parental classification system by medical domain. Its production, updates, and mappings to major biomedical resources rely on standardized and published procedures, continuous literature review, manual curation and expert validation, reflecting advancements in RD knowledge and clinical practice. Presented data metrics were computed using the Orphanet July 2025 release to quantitatively characterize the content, structure, classification, and semantic alignments of the Orphanet Nomenclature and Classification system.

**Results:** As of July 2025, the Orphanet Nomenclature of RD includes a total of 9,784 active clinical entities, including 6,527 disorders (corresponding to the RD definition), 1,084 subtypes of disorders, and 2,173 groups of disorders. Disorders are multiclassified into 29 classification hierarchies, each corresponding to a distinct medical domain, accurately representing the complex multisystemic nature of RD. Extensive qualified mappings ensure semantic interoperability: 97.4% of disorders are mapped to at least one ICD-10 code (6.4% with an exact proximity relationship), 71.8% are mapped to at least one ICD-11 MMS code (14.7% with an exact relationship) and 94.8% are mapped to SNOMED CT (all with an exact relationship). Genetic disorders represent 72.2% of all RD, and 63.4% are mapped to at least one phenotypic OMIM number.

**Conclusions:** The Orphanet Nomenclature and Classification of RD is the only RD-specific interoperable medical terminology meeting the needs of healthcare, research, and public health systems. By addressing the underrepresentation of RD in medical terminologies, it enables accurate RD identification, coding, and monitoring, supporting cross-border data interoperability, and contributing to improved knowledge, policy-making, and ultimately better care for people living with a RD.

## Introduction

Although rare diseases (RD) are individually uncommon, with over 6,500 different known RD, they collectively affect an estimated 329-624 million people worldwide [1,2]. Despite this significant global burden, available data indicate that 85% of RD affect less than 1 person per million [1]. As a result, the critical amount of data necessary to improve knowledge, care, and treatment can only be achieved through cumulative data collection across different countries in a standardized manner [3,4]. Cross-border data collection, sharing, and exploitation to support evidence-based policy making, healthcare, and research are therefore essential in the field of RD, as underscored by numerous international and European initiatives [5].

Over the past decades, numerous definitions of RD have been proposed, reflecting local legislations and public health priorities [4,6–11], most of which use point prevalence as the common epidemiological indicator. However, the threshold for rarity varies geographically, ranging from less than 1 per 2,500 in Japan [12], to less than 50 per 100,000 in Europe [13], or no more than 200,000 persons in the United States [14]. Furthermore, definitions can differ clinically, sometimes including restrictions such as “life-threatening” or “chronically debilitating” [6,13,15]. To address these inconsistencies, Rare Diseases International (RDI), in collaboration with a global panel of experts and the World Health Organization (WHO), has developed an Operational Description of Rare Diseases, defining a RD as a “medical condition with a specific pattern of clinical signs, symptoms, and findings that affect fewer than or equal to 1 in 2,000 persons living in any World Health Organization-defined region of the world”. [16]. Despite these efforts, the lack of harmonization among countries continues to impact RD research, diagnosis, and healthcare planning [3,5,8].

To take public health action on RD, it is fundamental to measure how many people are affected by these diseases in a defined population (prevalence), how these conditions impact those affected, and how to monitor their medical and societal consequences. [1,4,17]. In this regard, the United Nations Resolution on People Living with a Rare Disease (PLWRD) “encourages Member States and relevant United Nations agencies to collect, analyze, and disseminate disaggregated data on persons living with a rare disease… to assess progress towards the improvement of the status of persons living with a rare disease”. This further highlights that adequate coding of RD patient data within health information systems is a prerequisite for an effective response to this global public health priority [18].

However, RD are under-represented in medical terminologies and classification systems used within health systems, with only a small fraction of RD possessing specific and unambiguous codes [19–22]. This limitation makes it difficult to trace RD patients in health information systems, preventing a robust understanding of the actual number of PLWRD. Only a small proportion of RD have a specific code in the 10^th^ edition of the International Classification of Diseases (ICD-10) [20–22], the most widely used codification system worldwide; the representation of RD has increased in the 11^th^ revision of the International Classification of Diseases (ICD-11) [21,23] and the Systematized Nomenclature of Medicine Clinical Terms (SNOMED CT) [24], though it still remains below the number of known RD. Moreover, RD cannot be easily identified in these reference systems, as they do not explicitly include any indication of the rarity of a disease. Finally, different regions use diverse semantic resources in their coding systems (e.g. ICD-10 or SNOMED CT), or diverse versions of the same resource (ICD local extensions, different ICD versions), hampering data sharing and interoperability. Given the already scarce nature of RD data, this barrier to data collection requires dedicated efforts to make RD recognizable in health information systems [25–27].

To tackle the challenges of RD codification and interoperability, Orphanet has developed and maintained a nomenclature of RD since 1997 and a classification of RD since 2006 [20,28]. Over time, adoption of the Orphanet Nomenclature of RD for RD codification within national health systems has been supported and endorsed by European and international policy initiatives aimed at improving RD identification and monitoring and RD cross-border data sharing. The Orphanet Nomenclature and Classification of RD is now recognized as the reference resource to support standardized RD codification across healthcare and research settings.

The aim of this paper is to present the Orphanet Nomenclature and Classification of RD, detailing its content as well as its production and update methodology, and an overview of its mappings to other medical terminologies and classification systems. Additionally, it provides a clear and up-to-date count of RD based on the above-mentioned consensus definition, and their distribution by medical domain. Finally, it discusses the importance of maintaining and adopting a standardized and reliable RD nomenclature and classification system to support clinical coding, research, healthcare interoperability, and policy-making.

## Methods

### Structural description of the Orphanet Nomenclature and Classification of RD

#### The core elements of the Orphanet Nomenclature and Classification of RD

The Orphanet Nomenclature of RD is a unique and multilingual standardized system aimed at providing a specific reference standard for RD, that is organized in a classification system, a multi-hierarchical and polyparental structure organized by medical domain according to diagnostic and therapeutic relevance. The Orphanet Nomenclature of RD is composed of clinical entities (a generic technical term used to describe the clinical items included in the nomenclature), each of which includes several elements to ensure precise and unambiguous representation of RD concepts (Figure 1) [29]:

- ORPHAcode: a unique and time-stable numerical identifier automatically assigned in ascending order to every entry registered in the Orphanet Nomenclature;
- Preferred term (or main name): the most widely accepted name for the diagnosis in the literature and/or in the medical community;
- Synonym(s): additional terms, including acronyms, fully equivalent to the preferred term, as many as necessary according to the information found in the literature;
- Classification level: indicates the level of precision of the clinical entity, among three possible granularity levels: Group of disorders, Disorder, or Subtype of disorder;
- Definition: a brief description of the major clinical characteristics that define the diagnosis and differentiate it from other related clinical entities.

**Figure 1.**
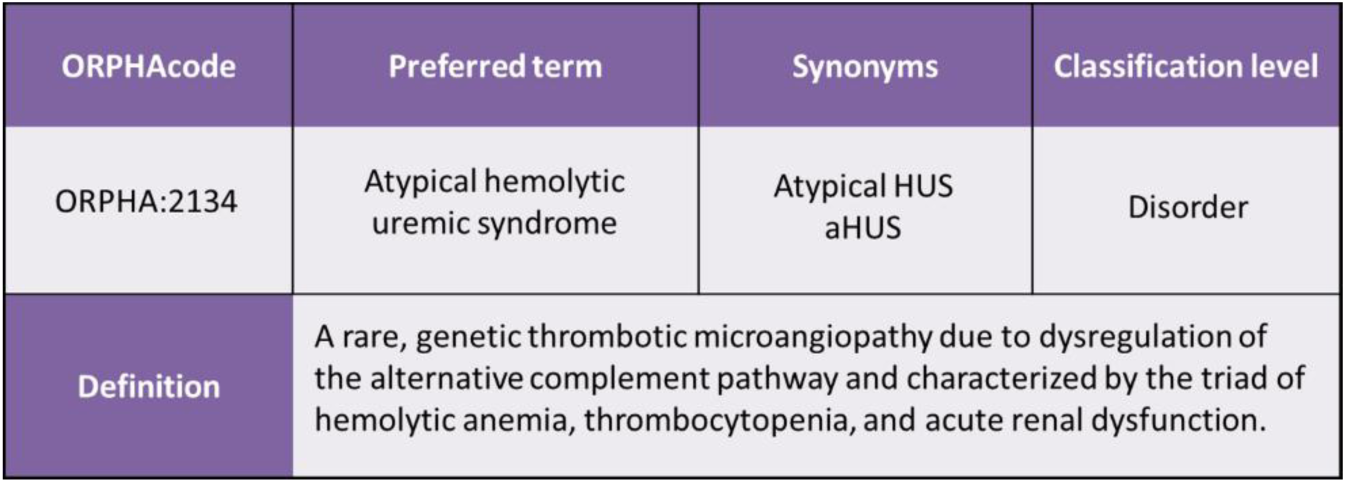
Technical elements composing the Orphanet Nomenclature of RD. Example of a clinical entity in the Orphanet Nomenclature of RD, defined by an ORPHAcode, a preferred term, as many synonyms as needed, a classification level, and a textual definition.

#### Naming rules and multilingual translation of the Orphanet Nomenclature of RD

In the absence of an international consensus on how to name diseases, preferred terms and synonyms are assigned according to standard naming rules developed by Orphanet to create a comprehensive RD nomenclature that aligns with current knowledge and clinical practice [30]. These rules ensure consistency across different disorder groups and medical domains, facilitating better integration and use of RD-related clinical information in databases and healthcare systems. The Orphanet Nomenclature of RD is produced in English (EN) and entirely translated into eight languages, French (FR), German (DE), Spanish (ES), Italian (IT), Dutch (NL), Portuguese (PT), Polish (PL) and Czech (CS), by translation teams located in member countries of the Orphanet Network. All translations are validated by RD expert medical doctors in each country. The English nomenclature serves as the basis for the translation of concepts; however, the translation process considers the particularities and local practices specific to every language, in particular with regards to the number of synonyms [31].

#### Classification levels and typology

The Orphanet Classification system is composed of the three above-mentioned classification levels, namely Group of disorders, Disorder, and Subtype of disorder (Figure 2). The classification level serves as the basis for parent-child relationships between related clinical entities. These relationships structure the Orphanet Classification according to successive criteria that are relevant in the clinical setting [29]. Additionally, each clinical entity is attributed a typology, a characteristic that, within the classification level, conveys the nosological definition of the clinical entity. For Group of disorders, typologies include Clinical group and Category; for Disorders, they include Disease, Clinical syndrome, Biological anomaly, Malformation syndrome, Morphological anomaly, and Particular clinical situation in a disease or syndrome; and for Subtype of disorder, they include Clinical subtype, Etiological subtype, and Histopathological subtype (Figure 2, definitions in Additional file 4) [29]. Of note, while generally used interchangeably to accommodate the widespread use of the term “rare diseases”, the terms “disease” and “disorder” designate different concepts within the Orphanet Nomenclature of RD. The term “disorder” is used to refer to the classification level that includes the six different typologies mentioned above, while the term “disease” represents a specific typology and a narrower clinical concept within the “disorder” classification level (Figure 2, Additional file 4) [29]. In the Orphanet Nomenclature and Classification system, the “disorder” level corresponds to the WHO-RDI consensus definition of “rare disease” [16].

**Figure 2.**
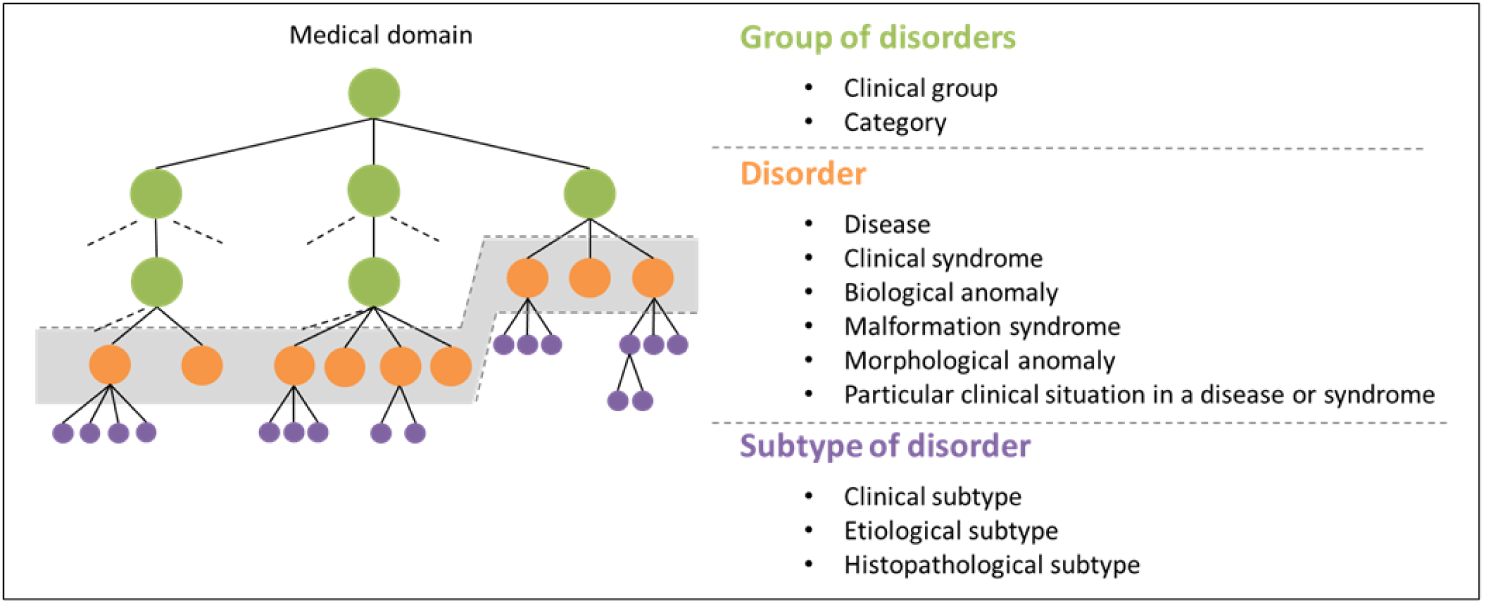
Classification levels and typologies of the Orphanet Nomenclature and Classification of RD. Every clinical entity in the Orphanet Nomenclature and Classification of RD is associated with a classification level conveying the precision of the diagnosis (Group of disorder, Disorder, or Subtype of disorder), and a unique typology within the classification level.

### Production and update process of the Orphanet Nomenclature and Classification of RD

#### Curation and update procedure of the Orphanet Nomenclature and Classification of RD

The Orphanet Nomenclature and Classification of RD is produced and updated according to a standardized and published procedure, in order to maintain an optimal quality of information, by a scientific team of information scientists at the Institut national de la santé et de la recherche médicale (Inserm) US14 service unit, Paris, France. The information provided in the Orphanet Nomenclature is sourced from up-to-date scientific and medical literature (more than 2,000 papers are consulted yearly), manually curated, reviewed and validated by Orphanet information scientists and RD medical experts, [29,32].

The maintenance of the Orphanet Nomenclature of RD is ensured through three main types of actions:

- Inclusion of a new clinical entity: to be included in the Orphanet Nomenclature, a disorder level clinical entity must correspond to a well-delineated clinical presentation, have a prevalence of fewer than or equal to 1 in 2,000 persons, and be described in at least two unrelated individuals in the literature, or two individuals from the same family when there is a clear genetic cause identified. Groups of disorders and subtypes of disorders are included according to established clinical practice and evolving medical knowledge. Before inclusion, the clinical entity is compared with existing entities to avoid overlaps and ensure classification consistency;
- Modification of an existing entity: a clinical entity in the Orphanet Nomenclature of RD can be modified to improve or update its information, including the preferred term, synonyms, classification level, typology or the definition. Clinical entities described before the genetic era, and lacking publications since then, are labeled as “Historic” and are kept in the Orphanet Nomenclature because their initial descriptions are still available and carry a potential for further characterization;
- Inactivation of a clinical entity: a clinical entity in Orphanet can be inactivated due to deprecation (it is now included in another concept), obsolescence (it has no valid reason to exist, such as being an imprecise or duplicate concept), or for no longer meeting the rare disorder prevalence criteria.

#### Management of inactivation, reactivation and replacement of ORPHAcodes

An ORPHAcode is permanently linked to the same clinical concept and is never deleted nor reused for a different entity, even if the original entity becomes inactive. In rare instances, an inactive clinical entity may be reactivated after reexamination with updated knowledge and reintroduced into the Orphanet Nomenclature and Classification with all relevant scientific information. To ensure continuity of information, especially in patient records and coding systems, Orphanet provides a replacement active clinical entity for deprecated and obsolete entities. This replacement code, or target code, must be used instead of the corresponding deprecated clinical entity as it aligns with the evolution of knowledge in the literature. For obsolete entities, the target often refers to a group of disorders, guiding users to explore the relevant classification to identify the most appropriate replacement code based on the patient’s diagnosis. Entities inactivated as non-rare in Europe are not given a replacement code, as they no longer fall under the RD scope.

### Classification of RD by medical domain

#### The Orphanet Classification hierarchies

The Orphanet Classification is structured in 34 major classification groups, individually referred to as classification hierarchy. Of these, 29 represent a distinct medical domain and are listed in Additional file 5. The remaining 5 hierarchies are ad-hoc classifications designed primarily for organizational purposes:

- Rare genetic diseases: this classification does not represent a distinct medical domain but is rather a derived genetic version that mirrors all the other clinical classification hierarchies.
- Rare systemic and rheumatological diseases of childhood: this classification is fully included in the Orphanet classification of rare systemic or rheumatologic disorders;
- Rare transplant-related diseases: this classification does not represent a medical domain but is used to classify disorders according to an intervention.
- Rare teratologic disorders: this classification is fully included in the Orphanet classification of rare developmental anomalies during embryogenesis.
- Rare disorder without a determined diagnosis after full investigation: this classification does not represent a medical domain and is used to classify one single disorder ORPHA:6616874, Rare disorder without a determined diagnosis after full investigation [33]. This is a code that does not conceptually fall under any of the other Orphanet classification hierarchies, as it allows for the identification in healthcare information systems of patients who remain undiagnosed after a full investigation.

Every classification consists of a tree structure starting with a unique ultimate parent group at the top representing the broadest clinical concept/medical domain (head of classification), then branching out into as many intermediate groups as necessary with increasing precision of clinical concepts, until reaching the first level of the confirmed diagnosis, the disorder. For any given disorder, subtypes can be added when this level of precision is required in clinical practice for appropriate management and follow-up.

#### Multi-hierarchical classification and linearization

Every clinical entity in the Orphanet Nomenclature of RD is introduced in as many classification hierarchies as necessary according to major medical specialties involved in the management of the disorder. This results in a multi-hierarchical and multi-parental classification structure that reflects the multi-dimensionality and complexity of RD (Figure 3). However, only one ultimate parent group is chosen as the entity’s preferential classification (preferential parent), according to the most severely affected body system, the most determining involvement for the prognosis, and/or the medical specialty that will most likely be relied on for the management of the disorder. This process, named linearization, enables coherent statistical analysis. For a substantial number of RD, no firm scientific basis exists to support assignment to a single medical domain, making preferential parent selection inherently complex. Linearization in these situations is a conventional framework guided by a set of published rules aimed at limiting arbitrariness and ensuring consistent application [34].

**Figure 3.**
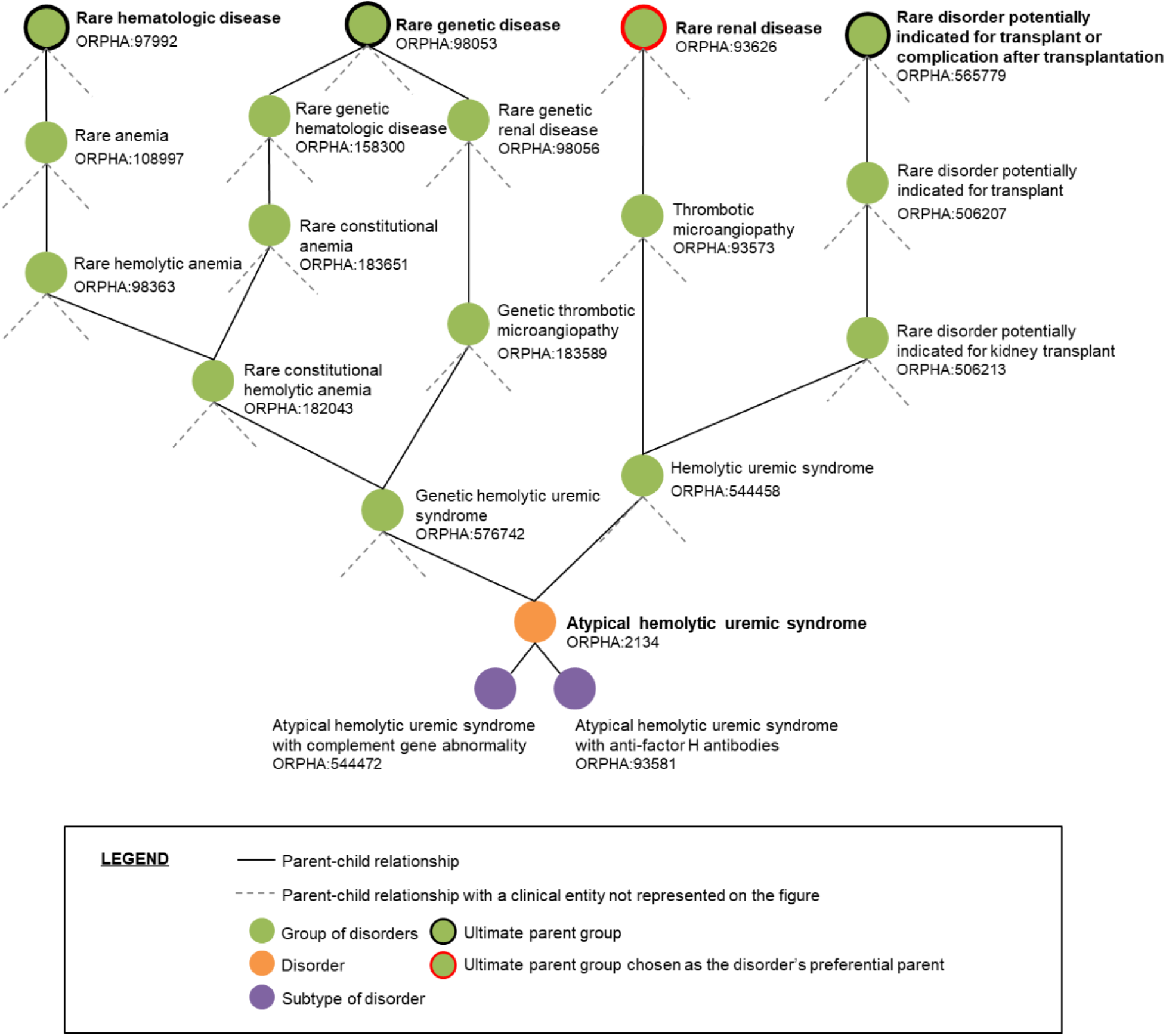
Representation of a disorder in the Orphanet Classification system. The disorder Atypical Hemolytic Syndrome (ORPHA:2134) is classified into four classification hierarchies. It has two direct parent groups, each with broader parent groups, culminating at the top of the hierarchy, where each ultimate parent group represents a medical domain. The Rare Renal Disorder group (ORPHA:93626) was designated as the preferential parent of the disorder Atypical Hemolytic Syndrome (ORPHA:2134). Note that all groups in the figure have additional child entities not shown here.

#### Mappings to other semantic resources

The Orphanet Nomenclature of RD is mapped to other international terminologies, classification systems, controlled vocabularies, knowledge bases, and ontologies according to a standardized and published procedure, to enable interoperability between different information systems [35]. Target standards include the WHO classification systems ICD-10 [36] and ICD-11, both Mortality and Morbidity Statistics (MMS) and Foundation [37], the clinical terminology SNOMED CT [38,39], the Online Mendelian Inheritance in Man (OMIM) knowledge base [40], the metathesaurus Unified Medical Language System (UMLS) [41], the controlled vocabularies Medical Subject Headings (MeSH) [42] and Medical Dictionary for Regulatory Activities (MedDRA) [43], the NIH Genetic and Rare Diseases Information Center (GARD) knowledge base [44], and the Mondo Disease Ontology (Mondo) [45].

The mapping cardinality always goes from the ORPHAcode to a code of the target resource. All mappings are qualified by a proximity relationship that defines the nature of the correspondence between the two concepts. The adopted proximity relationships are:

- E (Exact): the Orphanet entity designated by an ORPHAcode and the target code have the same range of application; they describe the same pathological entity;
- BTNT (Broader Term to Narrow Term): the Orphanet entity designated by an ORPHAcode has a broader range than the target code;
- NTBT (Narrow Term to Broader Term): the Orphanet entity designated by an ORPHAcode has a narrower range than the target code.

Mappings are generally validated at the Orphanet disorder and subtype levels; however, when an exact corresponding concept exists in the target resource, mappings may also be retained at the group level. For ICD-10, ICD-11 and OMIM, mappings are maintained with all three possible proximity relationships, whereas for SNOMED CT, UMLS, MeSH, MedDRA, GARD, and Mondo, only exact matches are retained.

The mappings with the WHO ICD-10 and ICD-11 are further characterized by a specificity relationship, describing how the equivalent clinical concept is represented in ICD [35]. The relationships are defined as follows:

- Specific code: the clinical concept represented by the ORPHAcode corresponds to a code in the ICD-10 tabular list/ICD-11 MMS exclusively denoting the concept;
- Index term: the clinical concept represented by the ORPHAcode is listed as an entry term in the ICD-10 alphabetical index or in the ICD-11 Foundation, without being assigned a dedicated tabular list/MMS code;
- Inclusion term (ICD-10 only): the clinical concept represented by the ORPHAcode appears as an inclusion term under an ICD-10 term in the tabular list without a distinct code;
- Attributed code: in the absence of an explicit corresponding term in ICD-10 or ICD-11 for the clinical concept represented by the ORPHAcode, an ICD code is attributed by Orphanet as the closest available clinical concept.

All mappings are manually curated and validated by an Orphanet information scientist and a medical doctor before publication, unless explicitly marked as *Not yet validated* to indicate pending medical review. Mappings with SNOMED CT concepts result from a collaborative process with SNOMED International, in which Orphanet disorder concepts are first evaluated for inclusion in SNOMED CT by a SNOMED International terminologist, and the resulting exact mappings are subsequently validated jointly by SNOMED International and Orphanet before publication. Notably, only mappings to ICD-10, ICD-11 and OMIM are distributed together with the official Orphanet Nomenclature files for coding, while the entirety of the mappings are distributed through Orphadata Science.

### Data extraction and analysis

#### Datasets description

Data metrics were computed using the Orphanet Scientific Knowledge Files delivered on the Orphadata Science website [46]. These datasets, structured in an Extensible Markup Language (XML) file format and in JSON format, are released in July every year at the same time as the official Orphanet Nomenclature files for coding (Nomenclature Pack) [47]. For this study, we analyzed the following datasets of the July 2025 release:

- the “rare diseases and alignments with terminologies and databases” dataset (<ISO>_product1.xml) in English, Czech, Dutch, French, German, Italian, Polish, Portuguese and Spanish languages; version 1.3.42 / 4.1.8 [2025-03-03] released 24 Jun 2025;
- the “linearization of rare diseases” English dataset (en_product7.xml); version 1.3.42 / 4.1.8 [2025-03-03] released 24 Jun 2025;
- the “genes associated with rare diseases” English dataset (en_product6.xml); version 1.3.42 / 4.1.8 [2025-03-03] released 24 Jun 2025;
- The “classifications of rare diseases” English datasets (en_product3_<X>.xml); version 1.3.42 / 4.1.8 [2025-03-03] released 24 Jun 2025;

Moreover, the differential files of the Orphanet Nomenclature packs for coding (ORPHAnomenclature_diff_en.xlsx) released from 2020 to 2025 were used to manually retrieve the annual inclusion and inactivation of disorders, and the human readable Orphanet-SNOMED-CT mapping file (ORPHA-SNOMEDCT_Mapping_File_production.xlsx), July 2025 version, released in October 2025, was used to report on the Orphanet-SNOMED CT mappings.

#### Metrics calculation

Metrics were computed using Python 3.8 scripts made available as dedicated Jupyter Notebooks (.ipynb files) in a Binder/Github-ready repository (https://github.com/Orphanet/orphanet_nomenclature_binder). This repository provides a publicly accessible, reproducible, and interactive computational environment that enables on-demand calculation of metrics using Orphadata files. A total of eight scripts were developed to calculate the necessary metrics and were annotated to document the calculation methodology. Metrics not computable via scripting were calculated manually, as detailed below.

The distribution of active clinical entities by classification level and typology was calculated using the script “1_number_of_Orphanet_clinical_entities”. This script uses the “rare diseases and alignments with terminologies and databases” dataset in English, filters active and inactive clinical entities, and subsequently counts them by classification level and typology. The same dataset was used to manually calculate the number of historical clinical entities by filtering for the “Historical entity” annotation.

The distribution of synonyms by classification level in English, as well as the number of adapted synonyms in Czech, Dutch, French, German, Italian, Polish, Portuguese and Spanish was calculated with the script “2_production_of_the_Orphanet_RD_terminology_in_english_and_other_language”. This script uses the “rare diseases and alignments with terminologies and databases” dataset in all target languages and sequentially filters and counts annotations for active clinical entities, and retrieves and counts their synonyms, grouping them by classification level.

The number of translated preferred terms in Czech, Dutch, French, German, Italian, Polish, Portuguese and Spanish was calculated manually by comparing the preferred terms of active clinical entities in each target language with the corresponding English preferred terms in the “rare diseases and alignments with terminologies and databases”. Terms differing from the corresponding English term were considered translated; terms identical to the corresponding English term were considered untranslated, with the exception of acronyms and Latin terms, which were considered translated.

The coverage of definitions at the disorder level in each language was calculated using the script “8_number_of_Orphanet_RD_with_definition_in_all_languages”. This script uses the “rare diseases and alignments with terminologies and databases” dataset in all target languages, filters active clinical entities at the disorder classification level, and retrieves and counts the corresponding definitions.

The annual disorder inclusions and inactivations for the period 2020–2025 were manually retrieved from the differential files of the Orphanet Nomenclature packs for coding released between 2020 and 2025.

The number of disorders classified in more than one classification hierarchy versus those classified in a single classification hierarchy was calculated using the script “4_number_of_Orphanet_RD_monoclassified_vs_multiclassified” which uses the “classifications of rare diseases” datasets to select disorders and assess their inclusion in one or multiple hierarchies. The same datasets are exploited by the script “5_distribution_of_Orphanet_RD_in_Orphanet_classifications” to retrieve and count the number of disorders included in each classification hierarchy. Of the 34 classification hierarchies in the Orphanet classification system, only 29 were included in the analysis. The previously described 5 hierarchies not corresponding to distinct medical domains or representing organizational or non-clinical groupings, were excluded. The distribution of disorders by preferential parent was calculated using the script “6_distribution_of_Orphanet_RD_by_preferential_parent”. This script selects all active disorders from the “rare diseases and alignments with terminologies and databases” dataset and uses the “linearization of rare diseases” dataset to retrieve their preferential parent, subsequently counting the total number of disorders linearized for each classification hierarchy.

The proportion of genetic disorders was calculated using the script “3_proportion_of_genetic_versus_non_genetic_Orphanet_RD”. This script retrieves all active disorders from the “rare diseases and alignments with terminologies and databases” dataset and assesses their inclusion in the “genes associated with rare diseases” dataset and the “classifications of rare diseases - rare genetic diseases classification” dataset. Inclusion in either dataset was considered indicative of a genetic disorder.

Mappings to other semantic standards were retrieved using the script “7_alignment_between_Orphanet_RD_and_international_terminologies” for ICD-10, ICD-11, and OMIM, and manually for UMLS, MeSH, MedDRA, GARD, and Mondo. In both cases, the “rare diseases and alignments with terminologies and databases” dataset was filtered by target standard to retrieve mappings at the Orphanet disorder level and their relationships. Orphanet-SNOMED CT mappings were retrieved manually by filtering the human-readable Orphanet-SNOMED-CT mapping file for mappings at the disorder level.

## Results

### Quantitative characterization of the Orphanet Nomenclature and Classification of RD

#### Counting and distribution of active clinical entities in the Orphanet Nomenclature and Classification of RD by classification level and typology

As of July 2025, the Orphanet Nomenclature of RD contains a total of 9,784 active clinical entities, including 6,527 disorders corresponding to the consensus definition of “rare disease” [16] (among which is a specific disorder code for undiagnosed patients after full investigation [33]), 1,084 subtypes of disorders, and 2,173 groups of disorders. Among all active entities, 3.3% (325 disorders and 1 subtype) are considered historic, with no publications over the last 25 years. The distribution of all active clinical entities by their classification level and their typology is presented in Table 1 and in Additional file 1. Definitions of each typology are provided in Additional file 4.

**Table 1.**
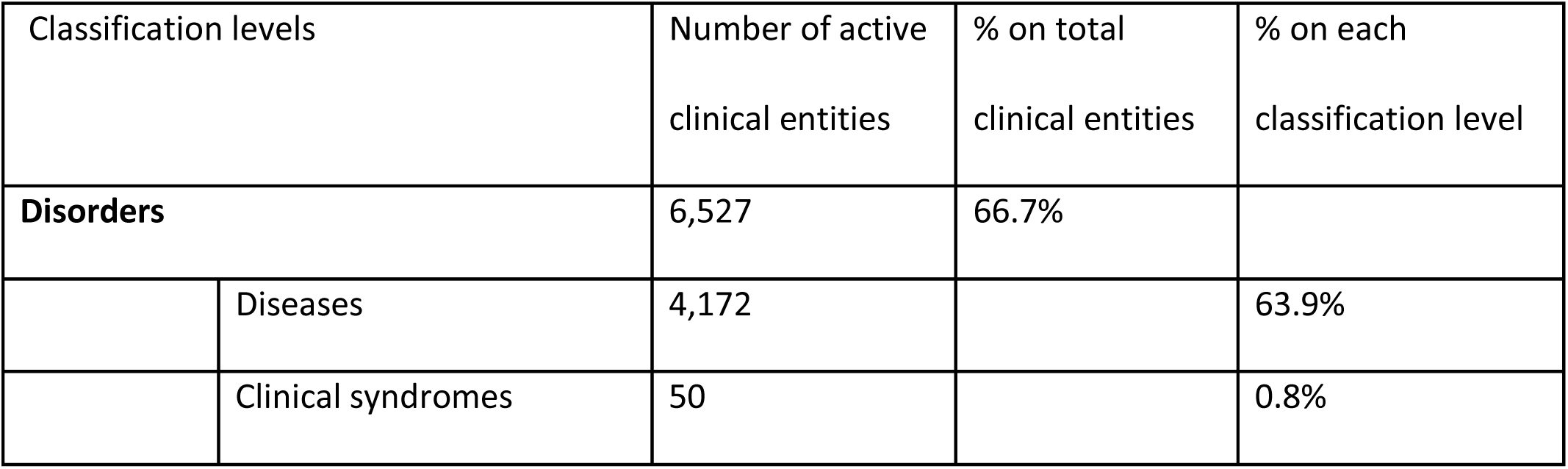
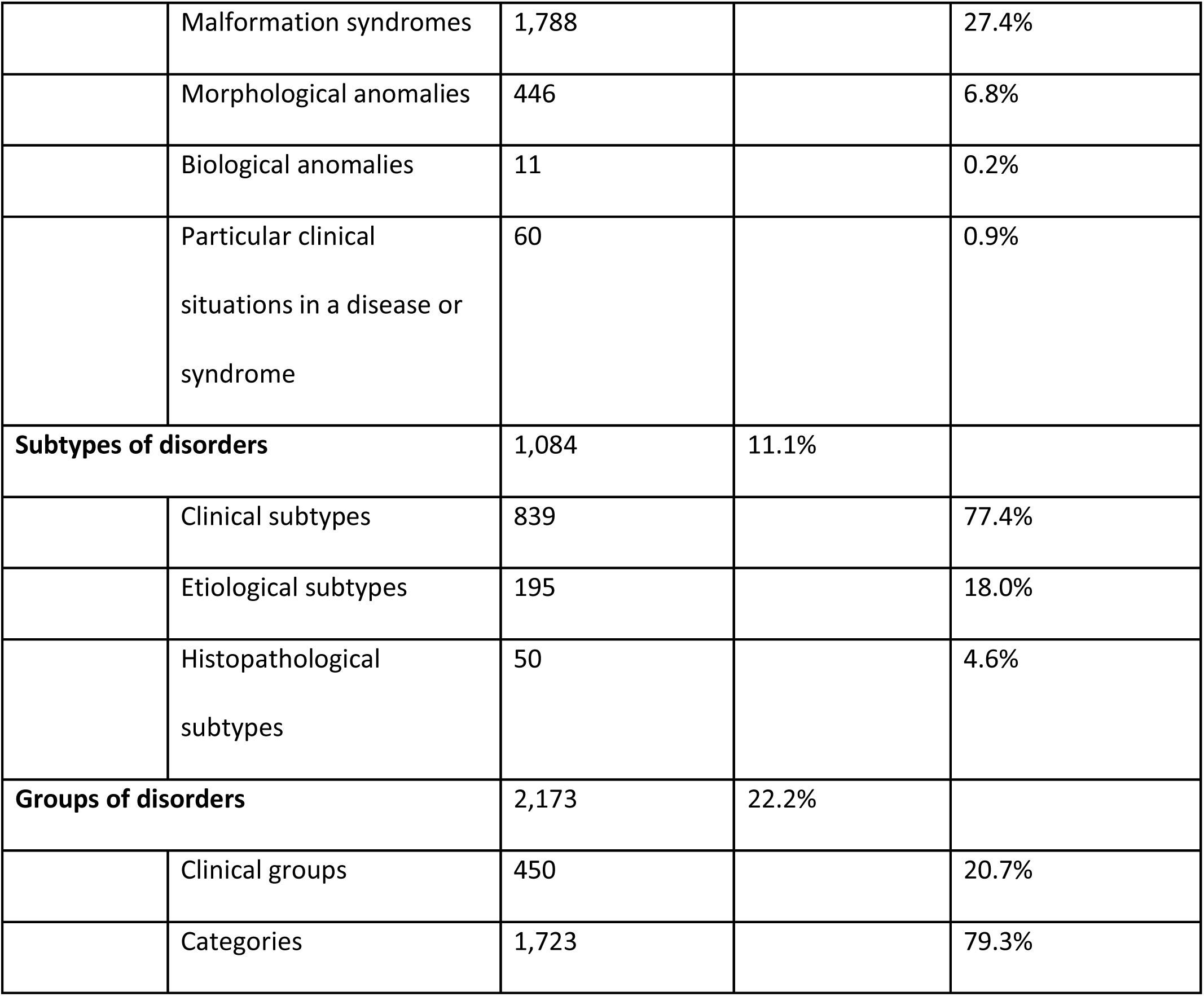
Distribution of active clinical entities in the Orphanet Nomenclature of RD by classification level and typology.

#### Counting and distribution of English synonyms across active clinical entities in the Orphanet Nomenclature of RD

Of the 9,784 active clinical entities in the Orphanet Nomenclature of RD, 6,311 (64.5%) carry at least one synonym in English, while the remaining 3,473 (35.5%) are not associated with any synonym. Of the 6,527 disorders, 4,805 (73.6%) carry at least one synonym in English, with a total number of synonyms at the disorder level amounting to 10,547. The minimum number of synonyms associated with a disorder is 1, and the maximum is 15, with a mean of 2.2 synonyms for disorders with at least one synonym. The complete distribution of synonyms by classification level is reported in Additional file 2.

#### Counting and distribution of translated preferred terms and synonyms across translation languages

All preferred terms associated with active clinical entities registered in the Orphanet Nomenclature of RD are translated in the following languages: French, German, Spanish, Italian, Dutch, Portuguese, Polish and Czech. Table 2 shows the rate of translation of preferred terms for every language. The very minor delay in translation is due to the different timings and frequencies at which translations are processed by every translation team. Non translated preferred terms are nevertheless included in English, pending translation. Synonyms can be adapted in all translated languages according to the medical practice in the target language; the number of adapted synonyms for every translation language is shown in Table 2. The complete distribution of synonyms by classification level in all translated languages is reported in Additional file 2. Additional translations are also available in Chinese (9,158 translated preferred terms, last update June 2020), Turkish (9,168 translated preferred terms, last update September 2021) and Ukrainian (9,612 translated preferred terms, last update July 2024), and are provided in separate files in the “Rare Diseases and Alignment with Terminologies and Databases” section of Orphadata Science [46].

**Table 2.** Number of active clinical entities with translated preferred term and adapted synonyms by language.

|  | FR | DE | ES | IT | NL | PT | PL | CS |
| --- | --- | --- | --- | --- | --- | --- | --- | --- |
| Number of translated preferred terms | 9,784 | 9,778 | 9,784 | 9,569 | 9,777 | 9,576 | 9,605 | 9,628 |
| % of translated preferred terms* | 100% | 99.9% | 100% | 97.8% | 99.9% | 97.9% | 98.2% | 98.4% |
| Number of adapted synonyms | 13,000 | 12,854 | 12,912 | 11,234 | 14,546 | 5,858 | 12,804 | 14,298 |

\*Percentages are calculated relative to the total number of active clinical entities with English preferred terms (n = 9,784).

#### Coverage and distribution of disorder definitions across languages

Disorder definitions are produced in English for all disorders included in the Orphanet Nomenclature of RD and are subsequently translated into the eight translation languages: French, German, Spanish, Italian, Dutch, Portuguese, Polish and Czech. Table 3 presents the coverage of disorder definitions at the disorder level in each language. Currently, approximately 94% of rare disorders have a definition in English, while the rate of coverage in other languages depends on the priorities defined by each translating country.

**Table 3.** Number of disorders with available definitions per language.

|  | EN | FR | DE | ES | IT | NL | PT | PL | CS |
| --- | --- | --- | --- | --- | --- | --- | --- | --- | --- |
| Number of disorders with a definition | 6,116 | 5,941 | 4,572 | 6,095 | 4,592 | 6,068 | 1,186 | 1,372 | 17 |
| % on the total number of disorders (n=6,527) | 93.7% | 91.0% | 70.1% | 93.4% | 70.4% | 92.9% | 18.2% | 21.0% | 0.26% |

#### Annual inclusion and inactivation of disorders in the Orphanet Nomenclature of RD over the last 6 years (2020-2025)

Since 2020, an average of 123 ± 72 (mean ± SD) new disorders per year have been added to the Orphanet Nomenclature, while an average of 44 ± 18 disorders per year have been inactivated. A detailed report of the number of newly included, obsoleted, deprecated and non-rare disorders for each year from 2020 to 2025 is provided in Table 4.

**Table 4.** Yearly overview of newly included and inactivated disorders in the Orphanet Nomenclature of RD (2020-2025).

|  | Newly included disorders | Obsoleted disorders | Deprecated disorders | Non-rare disorders |
| --- | --- | --- | --- | --- |
| 2020 | 74 | 35 | 24 | 5 |
| 2021 | 53 | 8 | 16 | 1 |
| 2022 | 92 | 29 | 5 | 4 |
| 2023 | 114 | 21 | 8 | 5 |
| 2024 | 253 | 38 | 22 | 9 |
| 2025 | 152 | 13 | 19 | 2 |

#### Distribution of rare disorders by medical domain in the Orphanet Classification of RD

The Orphanet Classification of RD has a multi-parental structure: 4,415 (67.7%) disorders are classified in more than one classification hierarchy while 2,106 (32.3%) are classified in just one classification, independently of the number of parents that they have in this same hierarchy. Figure 4 illustrates this multi-parental structure by comparing the number of disorders linearized to each medical domain and therefore counted once (gray bars), with the number of disorders classified in one or more medical domains and therefore counted as many times as they appear in each classification (white bars). As an example, while 1,096 disorders are primarily classified as neurological disorders, a total of 2,492 disorders present a major neurological involvement. Based on the linearization rules, the majority of rare disorders fall into the rare developmental defect during embryogenesis classification (32.3%, 2,109 disorders), closely followed by the rare neurologic disorders classification (16.8%, 1,096 disorders), amounting together to almost 50% of all rare disorders. The following most-represented classifications are the rare neoplastic disorders at 7.6%, the rare inborn errors of metabolism at 6.1%, the rare skin disorders at 6%, the rare bone disorders at 5.4%, the rare ophthalmic disorders at 3.7%, the rare immune disorders at 3.2%, the rare endocrine disorders at 3%, the rare hematologic disorders at 2.8%, the rare infectious disorders at 2.7%, and the rare systemic or rheumatologic diseases at 2.5%. The remaining classifications each include fewer than 100 linearized disorders each, collectively representing the residual 8% of rare disorders (Figure 4 gray bars; Additional file 3).

**Figure 4.**
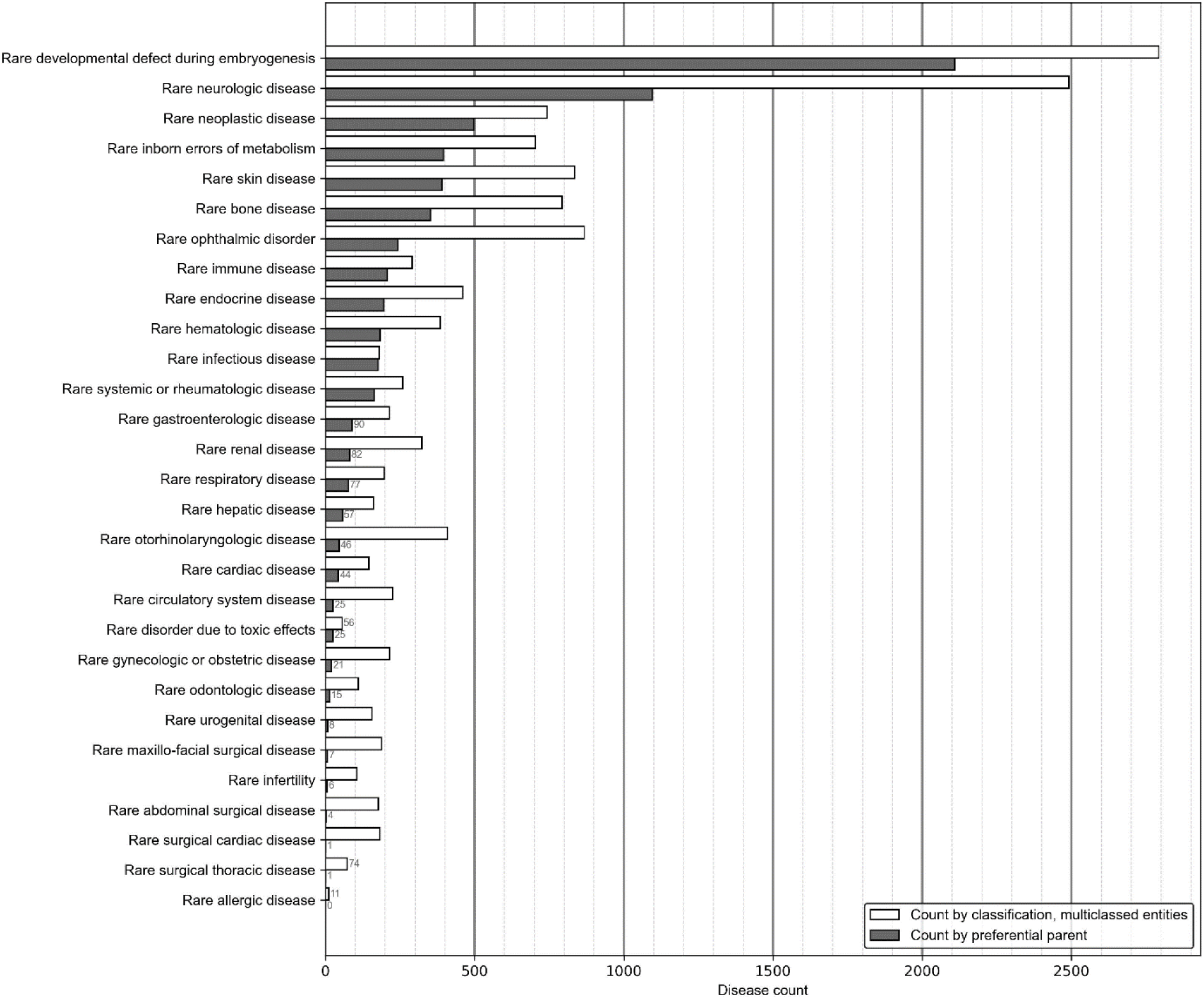
Distribution of rare disorders by medical domain according to their classification. Grey bars represent the number of disorders linearized according to their preferential parent classification, while white bars indicate the distribution of multi-classified disorders, counted as many times as they appear in the relevant classification hierarchies. Classifications containing fewer than 100 disorders are annotated with exact counts to enhance readability. The analysis includes 29 classification hierarchies corresponding to distinct medical domains; 5 hierarchies representing organizational or non-clinical groupings were excluded, as described in the Methods section. Six disorders were excluded from this analysis because they correspond to biological anomalies without clinical manifestations or were linearized under the classifications of rare genetic disorders or rare transplant-related disorders. ORPHA:616874 Rare disorder without a determined diagnosis after full investigation was also excluded.

A similar pattern is observed when considering multiclassified disorders, where the largest numbers are again found in the rare developmental defect during embryogenesis classification (2,793 disorders) and the rare neurologic disorders classification (2,492 disorders). The following classifications, however, contain considerably less multiclassified disorders and are, in descending order: rare ophthalmic disorders (868 disorders); rare skin disorders (835 disorders); rare bone disorders (793 disorders); rare neoplastic disorders (743 disorders); and rare inborn errors of metabolism (704 disorders). All remaining hierarchies contain less than 500 multiclassified disorders each, with only 3 hierarchies containing less than 100 (Figure 4 white bars; Additional file 3).

#### Coverage and proximity of the mappings between the Orphanet Nomenclature of RD and other semantic resources

Of the 6,527 active disorders registered in Orphanet, 6,355 (97.4%) are aligned to at least one code ICD-10, with only 415 (6.4% of all disorders) having an exact proximity relationship. The remaining aligned codes are either present in the ICD-10 alphabetical index/are included in the tabulated list under the ICD-10 code (655 disorders) or they have been attributed by Orphanet to the ICD-10 code that corresponds to the closest entity according to Orphanet’s rules (Table 5). The work on ICD-11 mapping has not been completed yet, however, as of July 2025, 4,683 disorders (71.8%) were mapped to at least one code in the ICD-11 MMS, with 958 (14.7% of all disorders) characterized by an exact proximity relationship and 3,013 (46.2%) present in the ICD11 Foundation with an exact uri. The remaining mappings have been attributed by Orphanet to the ICD-11 code corresponding to the closest entity (Table 5).

**Table 5.** Number of active disorders mapped to other semantic resources.

|  | Number of active disorders with at least one mapping | % on total disorders (n=6,527) | Number of active disorders with an exact proximity relationship | % on total disorders (n=6,527) |
| --- | --- | --- | --- | --- |
| ICD-10 | 6,355* | 97.4% | 415 | 6.4% |
| ICD-11 | 4,683* | 71.8% | 958 | 14.7% |
| SNOMED CT | 6,191 | 94.8% | 6,191 | 94.8% |
| OMIM | 4,141 | 63.4% | 3,319 | 50.9% |
| UMLS | 6,501 | 99.6% | 6,501 | 99.6% |
| MeSH | 2,735 | 41.9% | 2,735 | 41.9% |
| MedDRA | 1,455 | 22.3% | 1,455 | 22.3% |
| GARD | 3,279 | 50.2% | 3,279 | 50.2% |
| Mondo | 6,194 | 94.9% | 6,194 | 94.9% |

\* Counts include mappings attributed by Orphanet when the rare disorder concept was not present in the target resource.

According to the October 2025 release of the Orphanet-SNOMED CT mapping, 6,191 active Orphanet disorders (94.8%) are mapped to one SNOMED CT code, all with an exact proximity relationship (Table 5).

Among all active disorders present in the Orphanet Nomenclature, 72.2% (n= 4,715) are genetic disorders. A total of 4,141 Orphanet disorders are mapped to at least one phenotypic OMIM number and 3,319 mapped disorders have an exact proximity relationship with the corresponding phenotypic OMIM number (Table 5).

In addition, exact mappings to external semantic resources are distributed as follows: 6,501 (99.6%) for the UMLS Metathesaurus; 2,735 (41.9%) for MeSH; 1,455 (22.3%) for MedDRA; 3,279 (50.2%) for GARD; and 6,194 (94.9%) for Mondo. Of note, only mappings to ICD-10, ICD-11 and OMIM are distributed together with the official Orphanet Nomenclature files for coding, while the entirety of the mappings are distributed through Orphadata Science.

## Discussion

### Understanding the necessity of a RD-specific terminology

While collectively common, RD are numerous, clinically heterogeneous, and share challenges directly related to their individual rarity: persons living with each RD are few and dispersed; knowledge about each RD is limited and often inaccessible to non-specialist health professionals; and expertise is scarce and dispersed as well. RD are hard to identify in medical records since most are absent from medical terminologies in use in hospitals or not labeled as rare, rendering them invisible in health systems. This lack of identification results in inequalities in accessing a timely diagnosis and appropriate care, hinders data collection on RD natural history and their individual and societal burdens, and complicates patient recruitment for clinical research and therapy development [4,8,17,48].

The availability of adequate codes for each RD is a well-recognized need at the international level [25]. Improved codification for RD has been cited as a priority in the 2009 EU Council Recommendation on an action in the field of rare diseases [27], and more recently by the Rare2030 recommendations [49]. Having codes for each RD would help international, European, national, and local health authorities obtain better knowledge of healthcare pathways and of the impact of RD on both specialized (such as centers of expertise) and general health care services, while also supporting budget planning for health and social services [8,25,27]. Adequate coding is also necessary for identifying patients for clinical trials at both national and transnational levels. Finally, a large proportion of RD patients remain undiagnosed despite undergoing a full investigation, but this particular situation cannot be captured by the codification systems in use.

RD codification requires a nomenclature and classification system that meets the requirements of a standard medical terminology to ensure implementation in health information systems and in systematic research collections, such as patient registries. A trustworthy and reliable medical terminology must meet strict quality requirements: univocity and unambiguity of coded concepts, unrestricted management of synonyms, consistency of semantic relationships between concepts, multilingual support, accuracy of translations, correctness of semantic alignments, and relevant coverage of identified use cases. Additionally, terminology standards must adhere to rigorous and essential quality procedures, ensure sustainability and scalability to support the rapid development of knowledge, and guarantee long-term stability, preservation of the meaning of concepts from previous versions, and traceability of changes [50].

A key prerequisite for a RD terminology is to adopt a clear and operational definition of RD. Orphanet adopts a clinical definition that encompasses all RD, regardless of whether the etiology is known or not. This core concept ensures a terminology that can be adapted to different use cases, i.e. health systems with diverse diagnostic capabilities or different stages of the patient’s diagnostic journey. A RD in Orphanet is therefore defined as a clinical entity characterized by a set of well-delineated phenotypic abnormalities, described in at least two independent cases in the medical literature, and a consistent evolution, allowing for a definitive clinical diagnosis. The rarity of a disease is defined according to the European prevalence threshold (not more than 1 in 2,000 persons in the European population). This choice was made for historical reasons, as Orphanet was developed in Europe and co-funded by the European Commission (EC). However, this definition is consistent with those used in other geographical areas, which adopt either a similar or more restrictive cutoff, with the exception of the United States, where diseases with a higher prevalence are included [1,14].

Another limitation of this definition is that it includes infectious diseases which are rare in Europe, even if they are common in other parts of the world. To address this, some countries having adopted the Orphanet Nomenclature of RD outside the European space, like Argentina, have adapted this chapter to exclude infectious diseases that are common in their countries [51]. These limitations underline the need for consensus around a global operational definition of RD, a goal the WHO has worked towards in collaboration with RDI, with the participation of a multi-stakeholder, international group of experts including an Orphanet representative (A.R.) [16]. The results of this work support Orphanet’s adoption of a clinical definition of RD and may lead in the future to adaptations of the Orphanet RD lists for the different WHO regions. In the meantime, Orphanet collaborates with non-European countries to help adapt the Orphanet Nomenclature of RD to local needs, improving global RD interoperability.

### Implications of the Orphanet terminology structure, mappings and updates

The Orphanet Nomenclature and Classification of RD is, to the best of our knowledge, the only RD terminology clearly defining three levels of granularity: group of disorders, disorder and subtype of disorders, with the disorder level complying with the internationally agreed-upon definition of RD. This distinction has essential implications.

A first implication is that it allows to unambiguously count the number of existing RD (i.e. the disorder classification level), preventing overcounting due to the aggregation of heterogeneous granularity levels. As of today, many enumerations of RD circulate, some derived from semi-automated integration of different databases and ontologies (most frequently Orphanet and OMIM), sometimes resulting in estimates exceeding 10,000 RD [8,52,53]. However, these methods aggregate different levels of granularity without clearly stating which RD definition is adopted, rather reflecting the growing knowledge of disease mechanisms or treatable genetic variants than an actual increase in the number of RD. While this overall figure aligns closely with Orphanet’s total active clinical entities (9,784 as per July 2025 release), it is essential to recognize that superimposing different levels of granularity and/or different etiologies of the same RD can lead to counting the same disease multiple times. Using this broader count in public health discussions without clarification can lead to misinterpretations, potentially affecting policy decisions, budget allocation, and healthcare planning. For epidemiological and public health purposes, the count of clinically defined RD (6,527 according to the Orphanet July 2025 release) provides an appropriate common denominator for comparing countries and healthcare systems with varying technological and diagnostic capacities. It also permits to follow the evolution of therapy development and of healthcare resource allocation, including assessment of how many RD benefit from these measures over time and across geographies. Conversely, for precision medicine or molecular diagnosis, a more granular approach including subtypes (1,084 in the July 2025 release) or information on genetic variants collected in patient records may be needed.

A second implication is that defining the disorder granularity level allows the prevalence of RD to be established accurately and facilitates comparability across countries and continents. A clear RD definition therefore results in accuracy in counting RD and, consequently, in counting patients, which is essential for assessing the magnitude of RD as a public health issue. Furthermore, while allowing patient coding at the lowest granularity level (i.e. Subtypes of disorder), the classification hierarchies also allow data aggregation at the disorder level, thereby permitting to unambiguously establish the prevalence of RD. Notably, the Orphanet Nomenclature of RD is the only system which provides a code for undiagnosed RD patients who underwent all available diagnostic possibilities. This RD population has been estimated to represent up to 50% of all RD patients, based on the experience of RD expert centers [33]. Being able to code these patients at the disorder level makes it possible to include them in research pathways and undiagnosed programs, as recommended by IRDiRC’s goal #1 [54–56], and to include them in future global RD point prevalence estimations.

The three-level classification structure of the Orphanet Nomenclature and Classification of RD supports consistent semantic relationships between concepts, enabling effective use across various use cases and settings. While the disorder level serves as the primary reference level for data sharing among systems and countries, and for epidemiological purposes, groups of disorders allow data aggregation for use cases analysis, i.e. for public health decision-making such as healthcare organization (for example, designation of centers of expertise by RD area). Meanwhile, subtypes of disorders allow for fine-tuning of RD codification at the clinical, etiological and histopathological level, and can be used for applications in genetics, precision medicine, and other research and innovation purposes [57–63].

The Orphanet Classification system is also able to capture the multisystemic nature of RD, as clinical entities are organized into 29 distinct medical domains and classified as many times as needed according to their relevant domain of care. This is reflected in the distribution of RD across classification hierarchies: the majority of RD fall within the rare developmental defect during embryogenesis and rare neurologic disease classifications, together accounting for nearly 50% of all RD. However, almost 70% of RD are classified in more than one classification, reflecting their multisystem involvement. This approach ensures a more accurate representation of the complex nature of RD and of their linked resources, allowing to better orient patients and medical doctors towards appropriate information and specialized care [57–59]. At the same time, the distribution of entities across preferential parent classifications is uneven, with some domains encompassing a large proportion of RD and others including comparatively fewer entities. This distribution has implications for statistical analysis and data representation, as domains with fewer entities may appear less represented when preferential parent classifications are used for data aggregation. However, the multi-hierarchical structure allows flexible aggregation strategies that can be tailored to specific epidemiological or public health questions.

The field of RD is a rapidly evolving one: with the continuous advancement in genetic knowledge and the growing interest in personalized medicine, new RD are described each month. Additionally, the scope of previously identified RD can be refined, with medical concepts being either merged or divided based on new discoveries or the examination of larger numbers of patients. To ensure that the Orphanet Nomenclature and Classification of RD remain up to date with current scientific knowledge, Orphanet conducts a monthly systematized review of the literature and continuously collaborates with medical experts, particularly from the European Reference Networks (ERNs) and international RD learned societies [32,64]. Decisions on modifications to ORPHAcodes are made after an extensive review of the literature, and are validated by the Orphanet medical and scientific committee, following published standardized procedures [29]. Between 2020 and 2025, a total of 738 new RD were included in the Orphanet Nomenclature of RD, averaging about 120 RD per year. This number reflects not only newly described RD, but also the outcomes of numerous time-effective collaborative projects with experts, aimed at restructuring and updating the classification to incorporate revised medical concepts. At the same time, Orphanet also applies a strict inactivation policy to preserve the stability and univocity of ORPHAcodes. Inactivated codes are never reused for other entities, and a substitution code is always suggested to maintain traceability. While unusual, it is possible to reintroduce an inactivated ORPHAcode when new evidence emerges that justifies a reconsideration [29]. This has occurred in cases such as Systemic lupus erythematosus ORPHA:536 and Immunoglobulin A nephropathy ORPHA:34145, which did not initially meet RD prevalence criteria and are now considered RD. Taken together, the integration of new RD and the number of inactivated ones contribute to a small and relatively stable annual increase in the total number of RD, averaging fewer than 80 RD per year. A small proportion of active clinical entities are identified as historic, corresponding to RD for which no new patients have been reported in the medical literature for 25 years. These entities are retained in the nomenclature to preserve diagnoses that could potentially re-emerge and to support retrospective analyses, while their specific status limits the risk of confusion.

To further ensure the univocity and accessibility of concepts, Orphanet implements an unrestricted management of synonyms. Synonyms must cover the exact scope of the concept and are added as needed, without limitations in number [29]. This approach maximizes semantic coverage across different usages and data sources. Additionally, Orphanet provides a standardized, medically validated definition for each RD included in the Orphanet Nomenclature. These definitions consist of short texts describing the clinical features to unambiguously delineate the entity and distinguish it from other RD, and are intended to support healthcare professionals in assigning the correct ORPHAcode to a patient [29,65]. To further ensure international interoperability, the Orphanet Nomenclature and Classification of RD is also a multilingual resource. Translations are performed according to established procedures that include a final manual medical validation [31]. The different translations also include an unlimited number of synonyms that are tailored according to the relevance for each language. Taken together, these elements ensure semantic consistency and facilitate ORPHAcode integration into health and research information systems.

RD is a field in which the boundaries between diagnosis, healthcare, and research are often blurred, therefore maintaining semantic interoperability between health and research sectors is essential. The multiplicity of reference semantic standards used in healthcare, research and regulatory science reflects the fact that each serves a distinct purpose, adopting diverse levels of granularity, and encoding clinical concepts according to its own organizing principles. For example, ICD classifications are optimized for morbidity and mortality statistics, billing and reimbursement, and administrative reporting, which often results in broad disease groupings. In contrast, research-oriented resources such as OMIM organize information around gene-phenotype relationships, frequently splitting a single clinically defined disease into multiple molecular entries or, on the contrary, merging distinct clinical disorders related to the same gene. Consequently, semantic alignments between terminologies entail inherent complexity and need domain expertise, and must not be represented as simple one-to-one correspondences, but rather as structured relationships, as described in studies investigating RD terminology harmonization and mapping processes [66]. Mapping strategies must therefore explicitly document mapping cardinality and capture varying degrees of conceptual proximity and specificity, in order to preserve meaning across contexts and avoid misinterpretation. In addition, while automated approaches may assist in identifying candidate correspondences, the validation of RD mappings requires expert medical review to ensure that assigned relationships accurately reflect clinical meaning. This is reflected in the way ORPHAcodes are mapped to different resources, with defined relationships and medical validation. As of July 2025, Orphanet has aligned 97.4% of its active rare disorders to at least one ICD-10 code, with only 6.4% of all RD being exactly represented. The work on ICD-11 mapping is ongoing, but by July 2025, 71.8% of Orphanet’s RD were mapped to at least one ICD-11 MMS code, with 14.7% all RD being exactly represented. Additionally, the October 2025 release of the Orphanet-SNOMED CT mapping shows that 94.8% of Orphanet’s active RD are mapped to SNOMED CT, with all exhibiting an exact proximity relationship. Regarding genetic diseases, Orphanet classifies 4,715 RD (72.2% of active disorders) as genetic (defined as either caused by a known gene or having a clear familial inheritance pattern), with 4,141 RD mapped to one or more phenotypic OMIM number. It is interesting to note that many genetic RD are mapped to multiple OMIM numbers, reflecting the fact that Orphanet adopts a clinical definition of disease, whereas OMIM organizes entries around individual gene-phenotype relationships.

Orphanet is currently the only primary producer of a scientifically validated nomenclature and classification of RD. Other terminologies are increasingly integrating Orphanet concepts as a source of RD information, acknowledging the critical scientific contribution Orphanet makes to the RD community. Outstanding collaborations between Orphanet, SNOMED International and WHO ensure that the RD content of SNOMED CT and the ICD-11 continue to be improved by the inclusion of RD concepts as defined by Orphanet [23,24].

In parallel with mappings to major reference standards, the alignment of ORPHAcodes with other semantic resources, including UMLS, MeSH, MedDRA, GARD, and Mondo, supports a range of use cases such as literature indexing, pharmacovigilance, regulatory reporting, and knowledge integration. Overall, this extensive and accurate mapping system ensures consistency of transcoding across multiple platforms, further enhancing the utility of the Orphanet Nomenclature of RD in clinical, research, and regulatory contexts [60–63,67,68].

### Recognition and adoption of the Orphanet Nomenclature of RD

Since 1997, Orphanet has maintained through its publicly available website an inventory of RD to provide information on RD and their dedicated services across Europe [28]. This has evolved over time into a nomenclature and classification system of RD, that is today the only RD-specific standard meeting the high-quality requirements of a medical terminology [20].

In 2014, the EC Expert Group on Rare Diseases adopted a recommendation on improving RD codification [25]. In this document, EU Member States are encouraged to assess the feasibility of implementing ORPHAcodes at a national level and to include RD codification into their national RD plans/strategies. In 2017, the EC Steering Group on Promotion and Prevention (SGPP) recognized ORPHAcodes as a best practice [69]. Since then, ORPHAcodes have been included in the set of common data elements for Rare Diseases Registration, released by the EU RD platform on RD registration to enhance interoperability between registries [70]. The Orphanet Nomenclature of RD has also played a key role in implementing the EU 2011 Cross-Border Healthcare Directive in regards to RD. As a result, ORPHAcodes have been recommended for tracking RD diagnoses in the European Common Semantic Strategy (2019) [71] and the Guidelines on Patient Summary (Release 3.2, March 2022 and Release 3.4, November 2024) [72], and have been integrated into the Code Systems used in the MVC - My Health @ EU - eHealth Digital Service Infrastructure (eHDSI) [73,74]. The functional specifications developed by the X-eHealth project were instrumental for including ORPHAcodes in the minimum dataset required for the electronic Patient Summary [72,75]. Finally, the World Health Assembly (WHA) resolution “Rare diseases: a global health priority for equity and inclusion,” adopted on the 24th of May 2025, urges Member States to commit to “considering the implementation of ICD-11, and where appropriate, interoperable codification systems for RD such as the Orphanet Nomenclature of rare diseases, at their earliest possibility, and in accordance with their available resources, in order to enable the recording, reporting and monitoring of rare diseases at the national and international levels” [76].

To support the effective and sustainable implementation in health and research information systems, the Orphanet Nomenclature of RD (ORPHAcodes) is distributed under an open license (CC-BY 4.0) on different platforms depending on use: Orphadata Science for structured datasets intended for large scale analyses [46], and the Orphacode.org platform for coding resources for health information systems [47]. Each annual release (in July of each year) is versioned and accompanied by differential files to ensure traceability of changes and continuity of coding, as per the recommendation of the RD-ACTION working group for routine maintenance of codification [77]. In addition, a suite of technical tools is provided to facilitate implementation into diverse systems, promote accurate patient codification, and ensure continuous support by the community. Finally, the implementation of ORPHAcodes within information systems should be guided by established and evolving guidelines and best-practice recommendations, including those developed within the RD-CODE and the OD4RD/OD4RD2 project [33,78–80]. This supporting infrastructure ecosystem is essential to ensure evolution over time and respond to emerging use cases in a rapidly evolving field.

Establishing a seamless data ecosystem for RD based on the Orphanet Nomenclature and Classification of RD is a key step towards increasing the knowledge in a field that has long remained largely invisible within health systems, and ultimately improving the lives of PLWRD [49]. Indeed, accurate codification of RD patients enables the production of data for both primary and secondary use. Primary use includes, among others: recognition of the RD condition by non-expert health professionals; ensuring best care practices at all stages of the patient’s care pathway; and recognition of undiagnosed RD patients who might benefit from new discoveries, potentially leading to establishing a diagnosis. Secondary use of RD data includes accurately measuring the prevalence of the entire RD population and monitoring public health interventions to inform evidence-based decision-making; producing real-world evidence for research and therapeutic development; and facilitating recruitment for clinical research.

### Conclusions

The Orphanet Nomenclature and Classification of RD is the only RD-specific, high-quality standard, interoperable medical terminology meeting the needs of healthcare, research, and public health systems. By providing a clinically based and scientifically validated framework, it helps close the RD visibility gap in health and research information systems, enabling consistent RD identification and monitoring. Its wide adoption will support cross-border and cross-sectoral data interoperability, contributing to improved knowledge, policy-making, and ultimately better care for people living with a rare disease.

## Acknowledgements

We would like to thank the Orphanet translating members for their essential contribution to the translation of the Orphanet Nomenclature of RD, as well as the European Reference Networks (ERNs) and the Filières de santé maladies rares (FSMR) for their continued involvement in nomenclature and classification revisions, and in the production of textual content.

## Funding

The Orphanet Nomenclature and Classification system has been developed and maintained thanks to Inserm support since 2007 as well as European support since 2009 (*RD-PORTAL S12305098-2006119; RD-PORTAL2 S12324970-20091215; ORPHANET EUROPE JOINT ACTION 20102206 ; EUCERD Joint Action 2011 22 01; ORPHANET OPERATING GRANT 20133305; RD-ACTION JOINT ACTION 677024; ORPHANETWORK DIRECT GRANT 831390; RD-CODE 826607; OD4RD 101070531; OD4RD2 101110100*). The funders had no role in study design, data collection and analysis, decision to publish, or preparation of the manuscript.

## Ethics statement

This study did not involve human participants, patient-level data, or identifiable personal information. The analyses were conducted exclusively using data derived from published literature and publicly available sources curated within Orphanet. Consequently, approval from an Institutional Review Board (IRB) or equivalent ethics committee was not required, and informed consent was not applicable.

## Conflicts of Interest

The authors declare that they have no conflicts of interest.

## Data availability

The datasets supporting the conclusions of this article are publicly available. Orphanet provides open access to rare disease nomenclature, classification, genetic data, and mappings to other terminologies (https://www.orpha.net). Structured datasets for large-scale analysis are distributed via the Orphadata Science platform (https://sciences.orphadata.com/) and all previous versions are archived in a dedicated GitHub repository (https://github.com/Orphanet/Orphadata_aggregated). Additional resources include the Orphanet Nomenclature files for coding (Nomenclature Pack) distributed on the ORPHAcodes platform (https://www.orphacode.org/) and the Orphanet Rare Disease Ontology (ORDO, https://www.orphadata.com/ordo/). All formats are maintained and updated annually or bi-annually to ensure accuracy and accessibility.

## Authors’ contributions

AR, CL, AO, and HA conceived the study and contributed to the overall research goals and aims. CL, MDC, ADS, MF, EG, MCG, SM, MT, FS, VL, and DL curated and maintained the data. CL and DL carried out the formal analyses. AR, MH, and SM secured the funding. Methodology was developed by AR, CL, AO, HA, and VL. AR and CL coordinated and administered the project. DL developed the software. AR and MH provided supervision. Validation was performed by CL, DL, FL, and VL. CL and DL prepared the visualizations. CL, AR, AO, and HA drafted the initial manuscript. CL, AR, SM, CR, VL, and MH critically revised and edited the manuscript. All authors read and approved the final version of the manuscript.

## Abbreviations

BTNT: Broader Term to Narrow Term
CS: Czech
DE: German
EC: European Commission
EN: English
ERN: European Reference Network
ES: Spanish
EU: European Union
FR: French
FSMR: Filière de santé maladies rares
GARD: Genetic and Rare Diseases Information Center
ICD: International Classification of Diseases
IT: Italian
MeSH: Medical Subject Headings
MedDRA: Medical Dictionary for Regulatory Activities
MMS: Mortality and Morbidity Statistics
Mondo: Mondo Disease Ontology
NL: Dutch
NTBT: Narrow Term to Broader Term
OMIM: Online Mendelian Inheritance in Man
ORDO: Orphanet Rare Disease Ontology
PL: Polish
PLWRD: People Living with a Rare Disease
PT: Portuguese
RD: Rare Diseases
RDI: Rare Diseases International
SGPP: Steering Group on Promotion and Prevention
SNOMED CT: Systematized Nomenclature of Medicine Clinical Terms
UMLS: Unified Medical Language System
WHO: World Health Organization

## Multimedia ppendix

**Additional file 1 (Excel file .xlsx)**

**Distribution of Orphanet clinical entities by classification level and typology.** This file is the output of Notebook 1_number_of_Orphanet_clinical_entities.

**Additional file 2 (Excel file .xlsx)**

**Production of the Orphanet RD terminology in English and translation languages.** This file is the output of Notebook 2_production_of_the_Orphanet_RD_terminology_in_english_and_other_languages.

**Additional file 3 (Excel file .xlsx)**

**Distribution of disorders in Orphanet Classifications according to their preferential parent and multiclassified disorders.** This file combines the outputs of Notebook 5_distribution_of_Orphanet_RD_in_Orphanet_classifications and Notebook 6_distribution_of_Orphanet_RD_by_preferential_parent.

**Additional file 4 (PDF file)**

**Glossary of key concepts and definitions used in the Orphanet Nomenclature and Classification of Rare Diseases.**

**Additional file 5 (Word file .docx)**

**Overview of the 34 Orphanet classification hierarchies and their status as distinct medical domains or organizational classifications**

## Notes

### Competing Interest Statement

The authors have declared no competing interest.

### Summary of Updates

This version of the manuscript has been revised to improve the structure and organization of the manuscript, clarity, and readability. The Introduction and Discussion sections were reorganized to reduce redundancy and better distinguish background information from interpretation of results. The Methods section was expanded to provide clearer descriptions of the analytical approach and terminology framework. Terminology and key definitions used throughout the manuscript were also clarified to ensure consistent usage.

## References

1. Nguengang Wakap S, Lambert DM, Olry A, Rodwell C, Gueydan C, Lanneau V, Murphy D, Le Cam Y, Rath A. Estimating cumulative point prevalence of rare diseases: analysis of the Orphanet database. Eur J Hum Genet 2020 Feb;28(2):165–173. PMID:31527858

2. Orphanet, Inserm. Rare diseases in numbers. 2026 Feb. Report No.: 1. Available from: https://www.orpha.net/pdfs/orphacom/cahiers/docs/EN/OrphanetRDFactsheet.pdf [accessed Feb 24, 2026]

3. European Parliament: Directorate-General for Parliamentary Research Services, Schmiegelow K, Fehn K, Westman D, Clemente Pesudo I, Atanasovska D, Vucina Pedersen N. Addressing challenges to European multi-country collaboration models for rare diseases. European Parliament; 2024. Available from: https://data.europa.eu/doi/10.2861/51755 [accessed June 15, 2025]

4. Ferreira CR. The burden of rare diseases. Am J Med Genet A 2019 June;179(6):885–892. PMID:30883013

5. European Parliament: Directorate-General for Internal Policies of the Union, Kamphuis B, De Jongh T, Bastiaanssen V. Tackling rare diseases – Challenges, opportunities and gaps for action on rare diseases in the European Union. European Parliament; 2024. Available from: https://data.europa.eu/doi/10.2861/263904 [accessed June 15, 2025]

6. Richter T, Nestler-Parr S, Babela R, Khan ZM, Tesoro T, Molsen E, Hughes DA, International Society for Pharmacoeconomics and Outcomes Research Rare Disease Special Interest Group. Rare Disease Terminology and Definitions-A Systematic Global Review: Report of the ISPOR Rare Disease Special Interest Group. Value Health J Int Soc Pharmacoeconomics Outcomes Res 2015 Sept;18(6):906–914. PMID:26409619

7. Khosla N, Valdez R. A compilation of national plans, policies and government actions for rare diseases in 23 countries. Intractable Rare Dis Res 2018 Nov 30;7(4):213–222. PMID:30560012

8. Haendel M, Vasilevsky N, Unni D, Bologa C, Harris N, Rehm H, Hamosh A, Baynam G, Groza T, McMurry J, Dawkins H, Rath A, Thaxon C, Bocci G, Joachimiak MP, Köhler S, Robinson PN, Mungall C, Oprea TI. How many rare diseases are there? Nat Rev Drug Discov 2020 Feb;19(2):77–78. PMID:32020066

9. Chan AYL, Chan VKY, Olsson S, Fan M, Jit M, Gong M, Zhang S, Ge M, Pathadka S, Chung CCY, Chung BHY, Chui CSL, Chan EW, Wong GHY, Lum TY, Wong ICK, Ip P, Li X. Access and Unmet Needs of Orphan Drugs in 194 Countries and 6 Areas: A Global Policy Review With Content Analysis. Value Health Elsevier; 2020 Dec 1;23(12):1580–1591. PMID:33248513

10. Wainstock D, Katz A. Advancing rare disease policy in Latin America: a call to action. Lancet Reg Health Am 2023 Feb;18:100434. PMID:36844013

11. Abozaid GM, Kerr K, McKnight A, Al-Omar HA. Criteria to define rare diseases and orphan drugs: a systematic review protocol. BMJ Open 2022 July 29;12(7):e062126. PMID:35906057

12. Hayashi S, Umeda T. 35 years of Japanese policy on rare diseases. Lancet Lond Engl 2008 Sept 13;372(9642):889–890. PMID:18790305

13. European Parliament and Council of the European Union. Regulation (EC) No 141/2000 of the European Parliament and of the Council of 16 December 1999 on orphan medicinal products. Publications Office of the European Union; 2000 Jan. Available from: https://eur-lex.europa.eu/eli/reg/2000/141/oj/eng [accessed June 15, 2025]

14. United States Congress. Orphan Drug Act, 21 U.S.C. § 360bb(a)(2). United States Government Publishing Office; 2023. Available from: https://www.govinfo.gov/app/details/USCODE-2023-title21/USCODE-2023-title21-chap9-subchapV-partB-sec360bb [accessed Aug 3, 2025]

15. Abozaid GM, Kerr K, Alomary H, Al-Omar HA, McKnight A. Global insight into rare disease and orphan drug definitions: a systematic literature review. BMJ Open British Medical Journal Publishing Group; 2025 Jan 1;15(1):e086527. PMID:39863413

16. Wang CM, Whiting AH, Rath A, Anido R, Ardigò D, Baynam G, Dawkins H, Hamosh A, Le Cam Y, Malherbe H, Molster CM, Monaco L, Padilla CD, Pariser AR, Robinson PN, Rodwell C, Schaefer F, Weber S, Macchia F. Operational description of rare diseases: a reference to improve the recognition and visibility of rare diseases. Orphanet J Rare Dis 2024 Sept 11;19(1):334. PMID:39261914

17. Valdez R, Ouyang L, Bolen J. Public Health and Rare Diseases: Oxymoron No More. Prev Chronic Dis 2016 Jan 14;13:E05. PMID:26766846

18. United Nations General Assembly. Addressing the challenges of persons living with a rare disease and their families: resolution. United Nations; 2021. Available from: http://digitallibrary.un.org/record/3953832 [accessed June 15, 2025]

19. Jetté N, Quan H, Hemmelgarn B, Drosler S, Maass C, Moskal L, Paoin W, Sundararajan V, Gao S, Jakob R, Ustün B, Ghali WA, IMECCHI Investigators. The development, evolution, and modifications of ICD-10: challenges to the international comparability of morbidity data. Med Care 2010 Dec;48(12):1105–1110. PMID:20978452

20. Rath A, Olry A, Dhombres F, Brandt MM, Urbero B, Ayme S. Representation of rare diseases in health information systems: The orphanet approach to serve a wide range of end users. Hum Mutat 2012;33(5):803–808. PMID:22422702

21. Aymé S, Bellet B, Rath A. Rare diseases in ICD11: making rare diseases visible in health information systems through appropriate coding. Orphanet J Rare Dis 2015 Mar 26;10:35. PMID:25887186

22. Aymé S, Rath A, Bellet B. WHO International Classification of Diseases (ICD) Revision Process: incorporating rare diseases into the classification scheme: state of art. Orphanet J Rare Dis 2010 Oct 19;5(Suppl 1):P1. doi: 10.1186/1750-1172-5-S1-P1

23. World Health Organization. Rare diseases (frequently asked questions). WHO – Int Classif Dis. Available from: https://www.who.int/standards/classifications/frequently-asked-questions/rare-diseases [accessed June 15, 2025]

24. SNOMED International. INSERM and SNOMED International release SNOMED CT to Orphanet map supporting representation and use. SNOMED Int News. 2021. Available from: https://www.snomed.org/news/inserm-and-snomed-international-release-snomed-ct-to-orphanet-map-supporting-representation-and-use [accessed June 15, 2025]

25. Commission Expert Group on Rare Diseases (CEGRD), European Commission. Recommendation on ways to improve codification for rare diseases in health information systems. 2014 Nov. Available from: https://health.ec.europa.eu/publications/recommendation-ways-improve-codification-rare-diseases-health-information-systems_en [accessed June 15, 2025]

26. Choquet R, Maaroufi M, de Carrara A, Messiaen C, Luigi E, Landais P. A methodology for a minimum data set for rare diseases to support national centers of excellence for healthcare and research. J Am Med Inform Assoc JAMIA 2015 Jan;22(1):76–85. PMID:25038198

27. Council of the European Union. Council Recommendation of 8 June 2009 on an action in the field of rare diseases. Publications Office of the European Union; 2009. Available from: https://eur-lex.europa.eu/legal-content/EN/TXT/?uri=celex%3A32009H0703%2802%29 [accessed June 15, 2025]

28. Orphanet, Inserm. Orphanet – Knowledge on rare diseases and orphan drugs. Orphanet. Available from: http://www.orpha.net/ [accessed July 30, 2025]

29. Orphanet, Inserm. Procedural document: Orphanet nomenclature and classification of rare diseases. 2023 June. Report No.: 05. Available from: https://www.orpha.net/pdfs/orphacom/cahiers/docs/GB/eproc_disease_inventory_R1_Nom_Dis_EP_05.pdf [accessed Aug 3, 2025]

30. Orphanet, Inserm. Procedural document: Rare disease nomenclature in English. 2017 Apr. Report No.: 1. Available from: https://www.orpha.net/pdfs/orphacom/cahiers/docs/GB/eproc_Disease_naming_rules_in_English_PR_R1_Nom_01.pdf [accessed Aug 3, 2025]

31. Orphanet, Inserm. Procedural document: Orphanet Rare Diseases Nomenclature Production in National Language. 2017 Sept. Report No.: 1. Available from: https://www.orpha.net/pdfs/orphacom/cahiers/docs/GB/eproc_Rare_disease_Nomenclature_Production__national-language.pdf [accessed Aug 3, 2025]

32. Orphanet, Inserm. Procedural document: Collaboration with networks of expertise for the revision of the Orphanet nomenclature and classification of rare diseases. 2024 Sept. Report No.: 1.1. Available from: https://www.orpha.net/pdfs/orphacom/cahiers/docs/GB/eproc_Collaboration_networks_R1_Nom_Rev_Exp_EP_10.pdf [accessed Aug 3, 2025]

33. Angin C, Mazzucato M, Weber S, Kirch K, Abdel Khalek W, Ali H, Maiella S, Olry A, Jannot A-S, Rath A. Coding undiagnosed rare disease patients in health information systems: recommendations from the RD-CODE project. Orphanet J Rare Dis 2024 Jan 27;19:28. PMID:38280999

34. Orphanet, Inserm. Procedural document: Linearisation rules for rare diseases. 2014 Sept. Available from: https://www.orpha.net/pdfs/orphacom/cahiers/docs/GB/Orphanet_linearisation_rules.pdf [accessed Aug 3, 2025]

35. Orphanet, Inserm. Procedural document: Orphanet ICD-10 Coding Rules for Rare Diseases. 2023 June. Report No.: 3. Available from: https://www.orpha.net/pdfs/orphacom/cahiers/docs/GB/Orphanet_ICD10_coding_rules_R1_Nom_ICD_EP_07.pdf [accessed Aug 3, 2025]

36. World Health Organization. International statistical classification of diseases and related health problems. 10th revision (ICD-10). 2010 edition. Geneva: World Health Organization; Available from: https://icd.who.int/browse10/2010/en ISBN:978-92-4-154834-2

37. World Health Organization. ICD-11: International Classification of Diseases, 11th Revision. ICD-11 Mortal Morb Stat. Available from: https://icd.who.int/en [accessed June 15, 2025]

38. U.S. National Library of Medicine. Overview of SNOMED CT. US Natl Libr Med – Health IT. U.S. National Library of Medicine; 2014. Available from: https://www.nlm.nih.gov/healthit/snomedct/snomed_overview.html [accessed July 25, 2025]

39. SNOMED International. What is SNOMED CT. SNOMED Int. Available from: https://www.snomed.org/what-is-snomed-ct [accessed July 25, 2025]

40. Amberger JS, Bocchini CA, Schiettecatte F, Scott AF, Hamosh A. OMIM.org: Online Mendelian Inheritance in Man (OMIM®), an online catalog of human genes and genetic disorders. Nucleic Acids Res 2015 Jan 28;43(Database issue):D789–D798. PMID:25428349

41. Bodenreider O. The Unified Medical Language System (UMLS): integrating biomedical terminology. Nucleic Acids Res 2004 Jan 1;32(Database issue):D267–D270. PMID:14681409

42. Lipscomb CE. Medical Subject Headings (MeSH). Bull Med Libr Assoc 2000 July;88(3):265–266. PMID:10928714

43. Brown EG, Wood L, Wood S. The Medical Dictionary for Regulatory Activities (MedDRA). Drug Saf 1999 Feb 1;20(2):109–117. PMID:10082069

44. National Center for Advancing Translational Sciences (NCATS), National Institutes of Health. Genetic and Rare Diseases Information Center (GARD). Genet Rare Dis Inf Cent. Available from: https://rarediseases.info.nih.gov/ [accessed June 15, 2025]

45. Vasilevsky NA, Matentzoglu NA, Toro S, Flack JE, Hegde H, Unni DR, Alyea GF, Amberger JS, Babb L, Balhoff JP, Bingaman TI, Burns GA, Buske OJ, Callahan TJ, Carmody LC, Cordo PC, Chan LE, Chang GS, Christiaens SL, Daugherty LC, Dumontier M, Failla LE, Flowers MJ, Garrett HA, Goldstein JL, Gration D, Groza T, Hanauer M, Harris NL, Hilton JA, Himmelstein DS, Hoyt CT, Kane MS, Köhler S, Lagorce D, Lai A, Larralde M, Lock A, Santiago IL, Maglott DR, Malheiro AJ, Meldal BHM, Munoz-Torres MC, Nelson TH, Nicholas FW, Ochoa D, Olson DP, Oprea TI, Osumi-Sutherland D, Parkinson H, Pendlington ZM, Rath A, Rehm HL, Remennik L, Riggs ER, Roncaglia P, Ross JE, Shadbolt MF, Shefchek KA, Similuk MN, Sioutos N, Smedley D, Sparks R, Stefancsik R, Stephan R, Storm AL, Stupp D, Stupp GS, Sundaramurthi JC, Tammen I, Tay D, Thaxton CL, Valasek E, Valls-Margarit J, Wagner AH, Welter D, Whetzel PL, Whiteman LL, Wood V, Xu CH, Zankl A, Zhang XA, Chute CG, Robinson PN, Mungall CJ, Hamosh A, Haendel MA. Mondo: Unifying diseases for the world, by the world. medRxiv; 2022. p. 2022.04.13.22273750. doi: 10.1101/2022.04.13.22273750

46. Orphanet, Inserm. Orphadata Science – Orphanet datasets. Sci Orphadata. 2025. Available from: https://sciences.orphadata.com/ [accessed July 30, 2025]

47. Orphanet, Inserm. Orphanet Nomenclature Pack for Coding. ORPHAcodes. 2025. Available from: https://www.orphacode.org/pack-nomenclature/ [accessed July 25, 2025]

48. Schieppati A, Henter J-I, Daina E, Aperia A. Why rare diseases are an important medical and social issue. The Lancet Elsevier; 2008 June 14;371(9629):2039–2041. PMID:18555915

49. Kole A, Hedley V, et al. Recommendations from the Rare 2030 Foresight Study: the future of rare diseases starts today. EURORDIS-Rare Diseases Europe (Rare2030 project); 2021 Feb. Available from: https://download2.eurordis.org/rare2030/Rare2030_recommendations.pdf [accessed Aug 3, 2025]

50. Direction générale de l’offre de soins, France numérique santé (e-Santé) – Ministry of Health, France. Référentiels d’intéropérabilité sémantique : mise en euvre de terminologies de référence pour le secteur santé-social en France. Ministère des Solidarités et de la Santé, France; 2015. Available from: https://esante.gouv.fr/sites/default/files/media_entity/documents/20150504_etude_terminos_phase2_diagnostic.pdf [accessed June 15, 2025]

51. Ministerio de Salud de la Nación, Argentina. Listado de enfermedades poco frecuentes en Argentina. Argent – Salud. Available from: https://www.argentina.gob.ar/salud/pocofrecuentes/listado [accessed June 15, 2025]

52. Lamoreaux K, Lefebvre, S., Levine, D., Erler, W., Hume, T. The Power of Being Counted: A more accurate count of rare diseases and steps to getting counted. RARE-X Data Platform (RARE-X); 2022 May. Available from: https://rare-x.org/wp-content/uploads/2022/05/be-counted-052722-WEB.pdf [accessed Aug 3, 2025]

53. Smith CIE, Bergman P, Hagey DW. Estimating the number of diseases – the concept of rare, ultra-rare, and hyper-rare. iScience 2022 Aug 19;25(8):104698. PMID:35856030

54. International Rare Diseases Research Consortium (IRDiRC). IRDiRC Vision & Goals. IRDiRC Irdircorg. Available from: https://irdirc.org/about-us/vision-goals/ [accessed June 15, 2025]

55. Boycott KM, Rath A, Chong JX, Hartley T, Alkuraya FS, Baynam G, Brookes AJ, Brudno M, Carracedo A, den Dunnen JT, Dyke SOM, Estivill X, Goldblatt J, Gonthier C, Groft SC, Gut I, Hamosh A, Hieter P, Höhn S, Hurles ME, Kaufmann P, Knoppers BM, Krischer JP, Macek M, Matthijs G, Olry A, Parker S, Paschall J, Philippakis AA, Rehm HL, Robinson PN, Sham P-C, Stefanov R, Taruscio D, Unni D, Vanstone MR, Zhang F, Brunner H, Bamshad MJ, Lochmüller H. International Cooperation to Enable the Diagnosis of All Rare Genetic Diseases. Am J Hum Genet 2017 May 4;100(5):695–705. PMID:28475856

56. Lochmüller H, Le Cam Y, Jonker AH, Lau LP, Baynam G, Kaufmann P, Lasko P, Dawkins HJ, Austin CP, Boycott KM. ‘IRDiRC Recognized Resources’: a new mechanism to support scientists to conduct efficient, high-quality research for rare diseases. Eur J Hum Genet Nature Publishing Group; 2017 Feb;25(2):162–165. PMID:27782107

57. Mazzucato M, Pozza LVD, Facchin P, Angin C, Agius F, Cavero-Carbonell C, Corrochano V, Hanusova K, Kirch K, Lambert D, Lucano C, Maiella S, Panzaru M, Rusu C, Weber S, Zurriaga O, Zvolsky M, Rath A. ORPHAcodes use for the coding of rare diseases: comparison of the accuracy and cross country comparability. Orphanet J Rare Dis 2023 Sept 4;18:267. PMID:37667299

58. Mazzucato M, Visonà Dalla Pozza L, Minichiello C, Toto E, Vianello A, Facchin P. Estimating mortality in rare diseases using a population-based registry, 2002 through 2019. Orphanet J Rare Dis 2023 Nov 17;18:362. PMID:37978388

59. Pichon T, Messiaen C, Soussand L, Angin C, Sandrin A, Elarouci N, Jannot A-S, on behalf of the BNDMR infrastructure team. Overview of patients’ cohorts in the French National rare disease registry. Orphanet J Rare Dis 2023 July 3;18(1):176. PMID:37400917

60. Gunne E, McGarvey C, Hamilton K, Treacy E, Lambert DM, Lynch SA. A retrospective review of the contribution of rare diseases to paediatric mortality in Ireland. Orphanet J Rare Dis 2020 Nov 4;15(1):311. PMID:33148291

61. Rico J, Echevarría-González de Garibay LJ, García-López M, Guardiola-Vilarroig S, Maceda-Roldán LA, Zurriaga Ó, Cavero-Carbonell C. The interoperability between the Spanish version of the International Classification of Diseases and ORPHAcodes: towards better identification of rare diseases. Orphanet J Rare Dis 2021 Mar 9;16(1):121. PMID:33750434

62. Walker CE, Mahede T, Davis G, Miller LJ, Girschik J, Brameld K, Sun W, Rath A, Aymé S, Zubrick SR, Baynam GS, Molster C, Dawkins HJS, Weeramanthri TS. The collective impact of rare diseases in Western Australia: an estimate using a population-based cohort. Genet Med 2017 May;19(5):546–552. PMID:27657686

63. Scanlon P, Ridler G, Say G, Kellett M, Charlesworth J, Neil A, Dickinson JL, Burdon K, Jose M, Wallis M. Measuring the impact of rare diseases in Tasmania, Australia. Orphanet J Rare Dis 2024 Oct 28;19(1):399. PMID:39468681

64. European Commission (Directorate-General for Health and Food Safety). European Reference Networks. Eur Comm – Public Health. Available from: https://health.ec.europa.eu/rare-diseases-and-european-reference-networks/european-reference-networks_en [accessed July 25, 2025]

65. Orphanet, Inserm. Procedural document: Creation and update of disease summary texts for the Orphanet encyclopaedia for professionals. 2021 June. Report No.: 2. Available from: https://www.orpha.net/pdfs/orphacom/cahiers/docs/GB/Orphanet_DiseaseSummaryText_R1_Prod_sum_EP_02.pdf [accessed Aug 3, 2025]

66. Zoch M, Gierschner C, Weidner J, Lorenz S, Henke E, Reinecke I, Kallfelz M, Weber S, Thomas C, Sedlmayr M. ORPHAcode, ICD-10, SNOMED and Co.: Evaluation of terminology mappings in the field of rare diseases. Health Technol 2025 May 1;15(3):611–622. doi: 10.1007/s12553-025-00980-w

67. Chiu ATG, Chung CCY, Wong WHS, Lee SL, Chung BHY. Healthcare burden of rare diseases in Hong Kong – adopting ORPHAcodes in ICD-10 based healthcare administrative datasets. Orphanet J Rare Dis 2018 Aug 28;13:147. PMID:30153866

68. Navarrete-Opazo AA, Singh M, Tisdale A, Cutillo CM, Garrison SR. Can you hear us now? The impact of health-care utilization by rare disease patients in the United States. Genet Med 2021;23(11):2194–2201. PMID:34183788

69. Directorate-General for Health and Food Safety, European Commission. Rare diseases. Eur Comm – Public Health. 2025. Available from: https://health.ec.europa.eu/rare-diseases-and-european-reference-networks/rare-diseases_en [accessed June 15, 2025]

70. European Commission – Joint Research Centre (JRC). Set of Common Data Elements for Rare Diseases Registration. EU RD Platf Eur Comm Sci Hub. Available from: https://eu-rd-platform.jrc.ec.europa.eu/set-of-common-data-elements_en [accessed June 15, 2025]

71. eHAction — Joint Action supporting the eHealth Network. Common Semantic Strategy for Health in the European Union (eHAction Deliverable D8.2.2). European Commission; 2019 May. Available from: https://health.ec.europa.eu/document/download/92ac8823-19c4-4641-bf4d-5d9bf021a600_en [accessed Aug 3, 2025]

72. Directorate-General for Health and Food Safety. eHN Guideline on Patient Summary (Release 3.4). European Commission; 2024 Nov. Available from: https://health.ec.europa.eu/system/files/2023-10/ehn_guidelines_patientsummary_en.pdf

73. The European Health Data Space. Terms and Conditions of Use of the Code Systems used in the MVC. Available from: https://webgate.ec.europa.eu/fpfis/wikis/display/EHDSI/Terms+and+Conditions+of+Use+of+the+Code+Systems+used+in+the+MVC [accessed Feb 25, 2026]

74. EHDSI. Code Systems content managed by the Central Terminology Services. Available from: https://webgate.ec.europa.eu/fpfis/wikis/display/EHDSI/Code+Systems+content+managed+by+the+Central+Terminology+Services [accessed Feb 25, 2026]

75. X-eHealth (eXchanging electronic Health Records in a commom framework) Consortium. D5.6 – Refine PS functional specifications to account for eHN Guidelines and rare diseases. European Commission; 2022. Available from: https://www.x-ehealth.eu/wp-content/uploads/2022/11/D5.6-Refine-PS-functional-specifications-to-account-for-eHN-Guidelines-and-rare-diseases.pdf [accessed June 15, 2025]

76. Executive Board, World Health Organization. Rare diseases: a global health priority for equity and inclusion (EB156(15)). World Health Organization; 2025 Feb. Available from: https://apps.who.int/gb/ebwha/pdf_files/EB156/B156_(15)-en.pdf [accessed July 21, 2025]

77. Work Package 5, RD-ACTION Joint Action. Recommendation for routine maintenance of codification resources for rare diseases (RD-ACTION Deliverable 5.5). European Commission (Health Programme) / RD-ACTION; 2018 July. Available from: https://www.rd-action.eu/wp-content/uploads/2018/09/677024_DEL5.5_Recommendation-routine-maintenance-codification.pdf [accessed July 25, 2025]

78. RD-CODE. Specification and implementation manual of the Master file for statistical reporting with ORPHAcodes. Available from: https://www.rd-code.eu/wp-content/uploads/2022/01/826607_D5-4_Specification-and-implementation-manual-of-the-Master-file-for-statistical-reporting-with-ORPHAcodes.pdf [accessed Feb 25, 2026]

79. RD-CODE. Standard procedure and guide for coding with Orphacodes. Available from: https://www.rd-code.eu/wp-content/uploads/2022/01/826607_D5-4_Standard-procedure-and-guide-for-coding-with-Orphacodes_final.pdf [accessed Feb 25, 2026]

80. OD4RD2. Semantic interoperability of data on rare diseases - ORPHAcodes as part of the coding system landscape. Available from: https://od4rd.eu/communication-material/OD4RD2%20MS%2015%20White%20Paper_final-V02.pdf [accessed Feb 25, 2026]

