## Additional_file_5 for "The Orphanet Nomenclature and Classification of rare diseases: a standard terminology for improved patient recognition and data interoperability"

**Overview of the 34 Orphanet classification hierarchies and their status as distinct medical domains or organizational classifications**

| Classification hierarchy | Type of hierarchy |
| --- | --- |
| Rare abdominal surgical diseases | Distinct medical domain |
| Rare allergic disease | Distinct medical domain |
| Rare bone diseases | Distinct medical domain |
| Rare cardiac diseases | Distinct medical domain |
| Rare cardiac malformations | Distinct medical domain |
| Rare circulatory system diseases | Distinct medical domain |
| Rare developmental anomalies during embryogenesis | Distinct medical domain |
| Rare diseases due to toxic effects | Distinct medical domain |
| Rare disorder without a determined diagnosis after full investigation | Ad-hoc classification for organizational purposes |
| Rare endocrine diseases | Distinct medical domain |
| Rare gastroenterological diseases | Distinct medical domain |
| Rare genetic diseases | Ad-hoc classification for organizational purposes |
| Rare gynaecological and obstetric diseases | Distinct medical domain |
| Rare haematological diseases | Distinct medical domain |
| Rare hepatic diseases | Distinct medical domain |
| Rare immunological diseases | Distinct medical domain |
| Rare inborn errors of metabolism | Distinct medical domain |
| Rare infectious diseases | Distinct medical domain |
| Rare infertility | Distinct medical domain |
| Rare neoplastic diseases | Distinct medical domain |
| Rare neurological diseases | Distinct medical domain |
| Rare odontological diseases | Distinct medical domain |
| Rare ophthalmic diseases | Distinct medical domain |
| Rare otorhinolaryngological diseases | Distinct medical domain |
| Rare renal diseases | Distinct medical domain |
| Rare respiratory diseases | Distinct medical domain |
| Rare skin diseases | Distinct medical domain |
| Rare surgical maxillo-facial diseases | Distinct medical domain |
| Rare surgical thoracic diseases | Distinct medical domain |
| Rare systemic and rheumatological diseases of childhood | Ad-hoc classification for organizational purposes |
| Rare systemic and rheumatological diseases | Distinct medical domain |
| Rare teratologic disorders | Ad-hoc classification for organizational purposes |
| Rare transplant-related diseases | Ad-hoc classification for organizational purposes |
| Rare urogenital diseases | Distinct medical domain |
