## Additional_file_4 for "The Orphanet Nomenclature and Classification of rare diseases: a standard terminology for improved patient recognition and data interoperability"

### Glossary of key concepts and definitions

**Biological anomaly**: a disorder defined by a set of physiological abnormalities without clearly associated clinical manifestations.

**Category**: a group of clinically heterogeneous disorders sharing one general feature, used to organize the classification.

**Classification level**: the position of a clinical entity within the hierarchical structure of the Orphanet classification, reflecting the degree of diagnostic precision assigned to that entity (Group of disorders, Disorder, or Subtype of a disorder).

**Clinical entity**: a generic technical term used to describe the clinical items included in the Orphanet Nomenclature of rare diseases (RD).

**Clinical group**: a group of clinically homogeneous disorders that share a similar etiology, course, outcome, and/or management.

**Clinical subtype**: subdivision of a disorder according to distinct clinical characteristics (severity, age of onset, particular clinical signs, etc.).

**Clinical syndrome**: a disorder with homogeneous therapeutic possibilities, regardless of the pathophysiological mechanism involved.

**Disease**: a disorder with homogeneous therapeutic possibilities and an identified pathophysiological mechanism. Developmental anomalies are excluded.

**Disorder**: a clinical entity characterized by a set of homogeneous phenotypic abnormalities and evolution allowing a definitive clinical diagnosis.

**Etiological subtype**: subdivision of a disorder according to distinct causes resulting in a similar clinical presentation.

**Group of disorders**: a collection of clinical entities sharing a set of common features.

**Head of classification**: the unique ultimate parent group at the top of the classification hierarchy, representing the broadest clinical concept or medical domain from which the entire hierarchical structure originates.

**Histopathological subtype**: subdivision of a disorder according to characteristic histological patterns.

**Malformation syndrome**: a disorder resulting from a developmental anomaly involving more than one morphogenetic field. Malformative sequences and associations are included.

**Morphological anomaly**: a disorder characterized by a morphological alteration resulting from a development anomaly involving a single morphogenetic field.

**Orphanet Classification system**: a multi-hierarchical and polyparental structure that defines relationships between the clinical entities included in the Orphanet Nomenclature of RD and organizes them into medical domains according to diagnostic and therapeutic relevance.

**Orphanet Nomenclature and Classification of rare diseases**: a multilingual, standardized, controlled medical terminology specific to RD, that integrates a structured naming system, the Orphanet Nomenclature of RD, with a hierarchical classification framework, the Orphanet Classification system.

**Orphanet Nomenclature of rare diseases**: a unique, multilingual, standardized system that provides specific semantic references for RD.

**Particular clinical situation in a disease or syndrome**: a set of phenotypic abnormalities presenting in a subset of patients under particular circumstances.

**Preferential parent**: the head of classification group to which a clinical entity is assigned in order to organize entities by medical specialty and to prevent multiple counting of entities that belong to more than one classification group, particularly for statistical analyses.

**Subtype of a disorder**: subdivision of a disorder according to a positive criterion.
